# SARS-CoV-2 infection in pregnancy is associated with robust inflammatory response at the maternal-fetal interface

**DOI:** 10.1101/2021.01.25.21250452

**Authors:** Alice Lu-Culligan, Arun R. Chavan, Pavithra Vijayakumar, Lina Irshaid, Edward M. Courchaine, Kristin M. Milano, Zhonghua Tang, Scott D. Pope, Eric Song, Chantal B.F. Vogels, William J. Lu-Culligan, Katherine H. Campbell, Arnau Casanovas-Massana, Santos Bermejo, Jessica M. Toothaker, Hannah J. Lee, Feimei Liu, Wade Schulz, John Fournier, M. Catherine Muenker, Adam J. Moore, Yale IMPACT Team, Liza Konnikova, Karla M. Neugebauer, Aaron Ring, Nathan D. Grubaugh, Albert I. Ko, Raffaella Morotti, Seth Guller, Harvey J. Kliman, Akiko Iwasaki, Shelli F. Farhadian

**Affiliations:** Department of Immunobiology, Yale School of Medicine, New Haven, CT, USA; Department of Obstetrics, Gynecology, and Reproductive Sciences, Yale School of Medicine, New Haven, CT, USA; Department of Pathology, Yale School of Medicine, New Haven, CT, USA; Department of Molecular Biophysics and Biochemistry, Yale University, New Haven, CT, USA; Department of Cell Biology, Yale School of Medicine, New Haven, CT, USA; Department of Epidemiology of Microbial Diseases, Yale School of Public Health, New Haven, CT, USA; Section of Pulmonary and Critical Care Medicine, Department of Medicine, Yale School of Medicine, New Haven, CT, USA; Department of Pediatrics, Yale School of Medicine, New Haven, CT, USA; Department of Immunology, University of Pittsburgh, Pittsburgh, PA, USA; Department of Laboratory Medicine, Yale School of Medicine, New Haven, CT, USA; Section of Infectious Diseases, Department of Medicine, Yale School of Medicine, New Haven, CT, USA; Department of Molecular, Cellular and Developmental Biology, New Haven, CT, USA; Howard Hughes Medical Institute, Chevy Chase, MD, USA

## Abstract

Pregnant women appear to be at increased risk for severe outcomes associated with COVID-19, but the pathophysiology underlying this increased morbidity and its potential impact on the developing fetus is not well understood. In this study of pregnant women with and without COVID-19, we assessed viral and immune dynamics at the placenta during maternal SARS-CoV-2 infection. Amongst uninfected women, ACE2 was detected by immunohistochemistry in syncytiotrophoblast cells of the normal placenta during early pregnancy but was rarely seen in healthy placentas at full term. Term placentas from women infected with SARS-CoV-2, however, displayed a significant increase in ACE2 levels. Using immortalized cell lines and primary isolated placental cells, we determined the vulnerability of various placental cell types to direct infection by SARS-CoV-2 *in vitro*. Yet, despite the susceptibility of placental cells to SARS-CoV-2 infection, viral RNA was detected in the placentas of only a subset (∼13%) of women in this cohort. Through single cell transcriptomic analyses, we found that the maternal-fetal interface of SARS-CoV-2-infected women exhibited markers associated with pregnancy complications, such as preeclampsia, and robust immune responses, including increased activation of placental NK and T cells and increased expression of interferon-related genes. Overall, this study suggests that SARS-CoV-2 is associated with immune activation at the maternal-fetal interface even in the absence of detectable local viral invasion. While this likely represents a protective mechanism shielding the placenta from infection, inflammatory changes in the placenta may also contribute to poor pregnancy outcomes and thus warrant further investigation.

## Introduction

Coronavirus disease 2019 (COVID-19), due to SARS-CoV-2 infection, is a public health emergency that has impacted the lives of millions of people around the world. The effect of SARS-CoV-2 infection in pregnant women is of particular concern: population-based studies suggest that pregnant women with COVID-19 are at increased risk for severe illness compared to non-pregnant women with COVID-19(1). SARS-CoV-2 infection during pregnancy has also been associated with increased risk of pregnancy complications such as preterm birth, premature rupture of membranes, and preeclampsia(2-4). However, the mechanisms underlying these poor outcomes are unknown, and their dependence on SARS-CoV-2 infection of the placenta remains poorly understood.

Studies of other coronaviruses suggest the potential for placental pathology during maternal coronavirus infection both through direct viral invasion at the placenta and through a secondary inflammatory reaction. Mouse hepatitis virus (MHV), a coronavirus of laboratory mice, infects placental cells *in vivo*(5), leading to placental inflammation and increased susceptibility to subsequent bacterial infection of the placenta(6). During the SARS pandemic of 2008, maternal infection with SARS-CoV-1 was associated with histological abnormalities but not with viral invasion of the placenta(7). Case reports during the current COVID-19 pandemic have demonstrated that SARS-CoV-2 is capable of infecting the placenta(8-10); however, the mechanism for viral entry remains unclear. While variable findings have been reported(11-15) multiple recent transcriptomic analyses of healthy placentas have suggested limited expression of the canonical SARS-CoV-2 receptor *ACE2* in the placenta and little to no co-expression of *ACE2* with its classical co-factor *TMPRSS2* at the transcriptional level(11-13). Thus, it remains unclear whether the placenta is susceptible to SARS-CoV-2 infection under normal physiological conditions or under conditions of systemic inflammation, such as that which occurs with maternal COVID-19. Moreover, it remains unknown whether placental pathology develops in the absence of viral infection of the placenta(10, 16, 17).

In this study, we investigated the susceptibility of the human placenta to SARS-CoV-2 infection over the course of pregnancy, through *in situ* analysis of ACE2 protein expression and through *in vitro* studies. Furthermore, we describe *in vivo* immune responses at the maternal-fetal interface in response to maternal SARS-CoV-2 infection during pregnancy.

## Results

### Clinical, virological, and histological features of COVID-19 cases

A total of 39 women were identified as positive for COVID-19 by SARS-CoV-2 reverse transcription quantitative PCR (RT-qPCR) via nasopharyngeal swab prior to or at the time of delivery hospitalization. Universal screening of women presenting to Labor and Birth at Yale New Haven Hospital began on April 2, 2020. Two women were found to be SARS-CoV-2 RT-qPCR-positive before the universal screening period began; both were symptomatic with pneumonia. During the universal screening period, an additional 37 were identified as SARS-CoV-2 positive. Twenty-two (56%) of the SARS-CoV-2-infected women had symptomatic COVID-19. There were five cases of severe COVID-19 disease, requiring the administration of supplemental oxygen or ICU stay. Thirty-eight of the 39 pregnancies resulted in live births, with a median Apgar score of 9 (range 4-9). Clinical and demographic information for the COVID-19 cases is presented in Table 1.

**Table 1.** Clinical characteristics of COVID-19 cases and histological controls.

|  | COVID-19 cases (n=39) | Subset of COVID-19 cases with placenta histology available (n=27) | Histological controls (n=10) | P |
| --- | --- | --- | --- | --- |
| **Maternal Age: Median (range)** | 32 (18 - 41) | 32.0 (18 - 41) | 31.5 (21 - 37) | 0.788 |
| **Gestational Age: Median (range)** | 38w6d (22w6d - 41w1d) | 39w0d (36w6d - 41w1d) | 39w1d (37w - 40w6d) | 0.650 |
| **Comorbidities** |  |  |  |  |
| *Hypertension* | 12 (31%) | 9 (33%) | 4 (40%) | 0.752 |
| *Preeclampsia* | 6 (15%) | 5 (19%) | 1 (10%) | 0.640 |
| *Diabetes\** | 10 (26%) | 7 (26%) | 2 (20%) | 0.932 |
| **Mode of delivery (C-section)** | 13 (33%) | 10 (37%) | 3 (30%) | 0.472 |
| **Neonatal Apgar (1 minute)** | Median: 9<br>Range: 4- 9 | Median: 9<br>Range: 4- 9 | Median: 9<br>Range: 4 - 9 |  |
| **BMI: Mean (St Dev)** | 33.0 (7.3) | 33.4 (7.7) | 27.1 (2.33) | **0.0194** |
| **COVID-19 features** |  |  |  |  |
| *COVID-19 symptoms at the time of delivery (%)* | 22 (56%) | 14 (52%) |  |  |
| *Maternal severe COVID-19\*\** | 5 (13%) | 3 (11%) |  |  |
| **Nasopharyngeal SARS-CoV-2 RT-qPCR** |  |  |  |  |
| *CT value\*\*\* : median (range)* | 33.2 (17.1 - 43.8)<br>(n=26) | 33.35 (17.1 - 43.8) (n=18) |  |  |
| *Positive SARS-CoV-2 RT-qPCR of placenta (n=15)* | 2/15 (13.33%) |  |  |  |

\*gestational or pre-gestational diabetes
\*\*ICU or need for supplemental oxygenation
\*\*\*N1 gene

Among the 15 placentas tested by RT-qPCR for the presence of SARS-CoV-2, viral RNA was detected in two (∼13%) of the placentas (Supplementary Table 1). One was from a 32-year-old woman who presented in labor at 38 weeks of gestation with symptoms of pneumonia, not requiring supplemental oxygen, who progressed to a healthy delivery. The neonate tested negative for SARS-CoV-2 by nasopharyngeal swab RT-qPCR at the time of delivery. The other was from a woman who presented at 22 weeks of gestation with severe COVID-19 pneumonia and developed preeclampsia and fetal demise, resulting in fetal loss at 22 weeks. This case was excluded from the placental analyses presented here, since the details of this case have been reported previously(10). Plasma from 12 SARS-CoV-2 infected women was tested for SARS-CoV-2 spike S1 protein-specific IgG and IgM antibodies (anti-S1-IgG and -IgM). No apparent differences in ELISA absorbance values were observed between symptomatic and asymptomatic infected mothers, or between pregnant and non-pregnant SARS-CoV-2 infected individuals (Supplementary Figure 1).

A total of 28 placentas from 27 COVID-19 mothers were available for histological analysis (one COVID-19 case resulted in the delivery of dizygotic twins). The COVID-19 placental cases available for examination did not differ from the overall cohort of COVID-19 pregnancies during the study period by maternal age, gestational age, mode of delivery, neonatal outcomes, or co-morbidities (Table 1). Placental specimens were examined by two independent pathologists blinded to the patient’s COVID-19 status, and were assessed for the presence of villitis, chorioamnionitis, intervillositis, for increased decidual lymphocytes, and for fetal and maternal vascular malperfusion. No significant differences were seen between cases and controls for these features (Supplementary Table 2). However, increased intervillous fibrin was seen in 33% of cases (9/27) but in none of the controls (Figure 1; p=0.036). We found no association between the presence of increased intervillous fibrin and clinical features, including the presence of COVID-19 symptoms, co-morbidities, mode of delivery, or BMI. Overall, our analysis suggests that increased intervillous fibrin may be the only distinct histologic feature observed in placentas from COVID-19 positive mothers.

**Figure 1.**
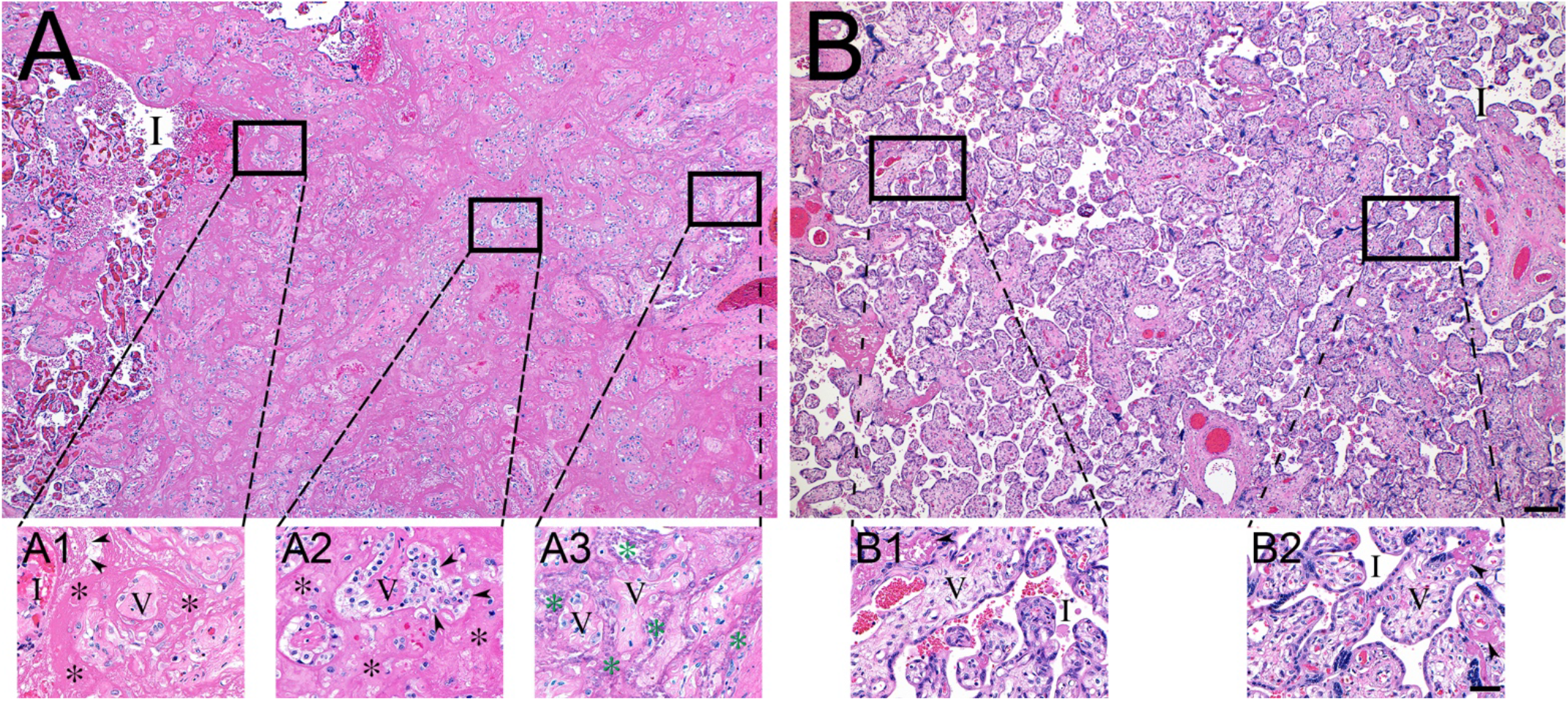
Histopathology of representative COVID-19 **(A)** and matched control **(B)** placentas. **(A)** COVID-19 placenta at low magnification revealed extensive intervillous fibrin deposition, with only occasional areas of open intervillous (I) spaces. **(A1)** High magnification at edge of blood filled intervillous space (I) and the earliest fibrin deposition (asterisks). Trapped chorionic villi (V) have become avascular and fibrotic. Initial fibrillar fibrin (arrow heads) can be seen at the blood-fibrin interface. **(A2)** Older area of intervillous fibrin (asterisks) and trapped villi (V) revealing migration of trophoblasts (arrow heads) into the fibrin matrix. **(A3)** Oldest area of intervillous fibrin became calcified (green asterisks), encasing villous remnants (V). **(B)** In sharp contrast, the control placenta revealed virtually no fibrin in the intervillous space (I). **(B1 and B2)** Representative magnified areas revealed normal villi (V) and open, maternal blood containing intervillous space (I), with only occasional foci of fibrin formation (arrow heads). Bars represents 200 µM for images A and B and 50 µM for images A1-B2.

### Decreased ACE2 protein expression in the placenta over the course of normal pregnancy

We assessed the potential for SARS-CoV-2 infection of the placenta by examining placental expression of ACE2, the canonical receptor required for SARS-CoV-2 infection. Prior transcriptome studies have suggested that *ACE2* is absent or expressed at low levels in placenta. Consistent with these previous reports, our analysis of bulk and single-cell RNA sequencing data in placenta from COVID-19 cases and controls demonstrates very low levels of *ACE2* gene expression at the term placenta (Supplementary Figure 2). However, when protein-level ACE2 expression was examined by immunohistochemistry, we found ACE2 to be highly expressed in syncytiotrophoblast cells in first and second trimester placentas, with ACE2 protein expression virtually absent in normal term placentas obtained from pre-pandemic controls (Figure 2B-F).

**Figure 2.**
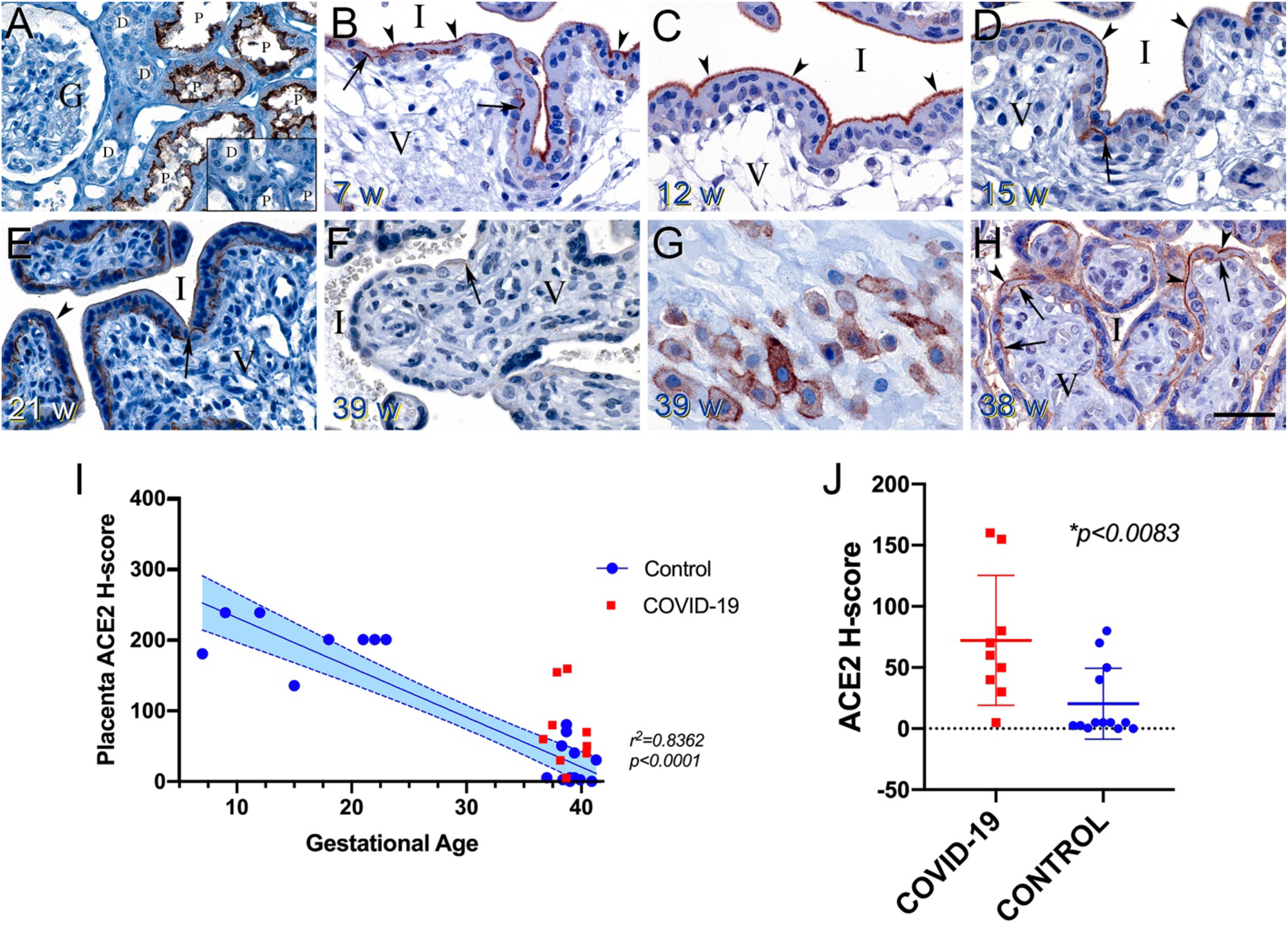
ACE2 protein expression in the placenta varies with gestational age. **(A)** Human kidney used as a positive control revealed strong apical staining of the proximal tubules (P). The distal tubules (D) and glomerulus (G) were negative. Inset shows a serial section of the same kidney stained with non-immune rabbit sera resulting in no staining. **(B-D)** Placentas derived from normal pregnancies between 7 and 15 weeks of gestation demonstrated strong, uniform, apical microvillus syncytiotrophoblast staining (arrow heads), and patchy strong basolateral staining at the cytotrophoblast–syncytiotrophoblast contact zone (arrows). Intervillous space (I) and villous core (V). **(E)** A normal 21-week placenta still exhibited syncytiotrophoblast surface staining (arrow head), but to a lesser extent than the earlier samples. Cytotrophoblast– syncytiotrophoblast contact zone staining was still prominent (arrow). **(F)** A representative normal placenta at 39 weeks revealed almost no ACE2 staining. Occasionally, staining at the cytotrophoblast–syncytiotrophoblast contact zone was noted (arrow) **(G)** Normal extravillous invasive trophoblasts from a 39-week placenta demonstrated strong surface expression of ACE2, with variable cytoplasmic staining. **(H)** Representative image of ACE2 expression in a 38-week placenta derived from a case of symptomatic maternal COVID-19. Reappearance of strong apical microvillus syncytiotrophoblast (arrow heads) and cytotrophoblast– syncytiotrophoblast contact zone staining (arrows) was observed. All sections were cut at 5 µM, except panel (E), which was cut at 10 µM. Bar represents 50 µM for images A-H. **(I)** ACE2 H-score demonstrated steady loss of placental ACE2 with increasing gestational age in healthy pregnancies (p<0.001). Linear regression (blue line) was fit to data from healthy controls (circles). 95% confidence interval is shown with dashed lines. Placentas derived from COVID-19 cases are depicted as red squares. **(J)** ACE2 H-score was significantly increased in term placentas from COVID-19 cases (squares) compared to uninfected, matched controls (circles).

While the expression pattern of ACE2 in the placenta decreased steadily over gestational age in placentas derived from healthy pregnancies (Figure 2I), we found that ACE2 protein was present at significantly higher levels in term placenta collected from COVID-19 cases (Figure 2J). These findings suggest that detection of *ACE2* mRNA expression is not a reliable surrogate for ACE2 protein expression in the placenta and, importantly, that ACE2-mediated risk for placental infection by SARS-CoV-2 may vary over the course of pregnancy, with our detection of higher ACE2 levels in the first and second trimesters suggesting the most vulnerability may exist prior to term.

### *In vitro* infection of primary isolated cytotrophoblasts by SARS-CoV-2

To determine whether the low rate of placental infection we observed in our case series was due to intrinsic resistance to SARS-CoV-2 infection by placental cells, we performed *in vitro* infections of placental cells to determine the infectious potential of SARS-CoV-2 at the placenta. We infected primary placental cells isolated from healthy term deliveries with a replication-competent infectious clone of SARS-CoV-2 expressing the fluorescent reporter mNeonGreen (icSARS-CoV-2-mNG)(18) for 24 hours.

We observed no detectable infection of primary isolated syncytiotrophoblasts (derived from cytotrophoblasts allowed to spontaneously differentiate for 72-96 hours), Hofbauer cells, or fibroblasts following 24 hours of infection. However, we found infection of primary isolated cytotrophoblasts, as observed by mNeonGreen reporter detection and staining for SARS-CoV-2 nucleocapsid (NP) (Figure 3A). These findings were consistent with SARS-CoV-2 N1 detection by RT-qPCR in cells infected at 24 hours (Figure 3B).

**Figure 3.**
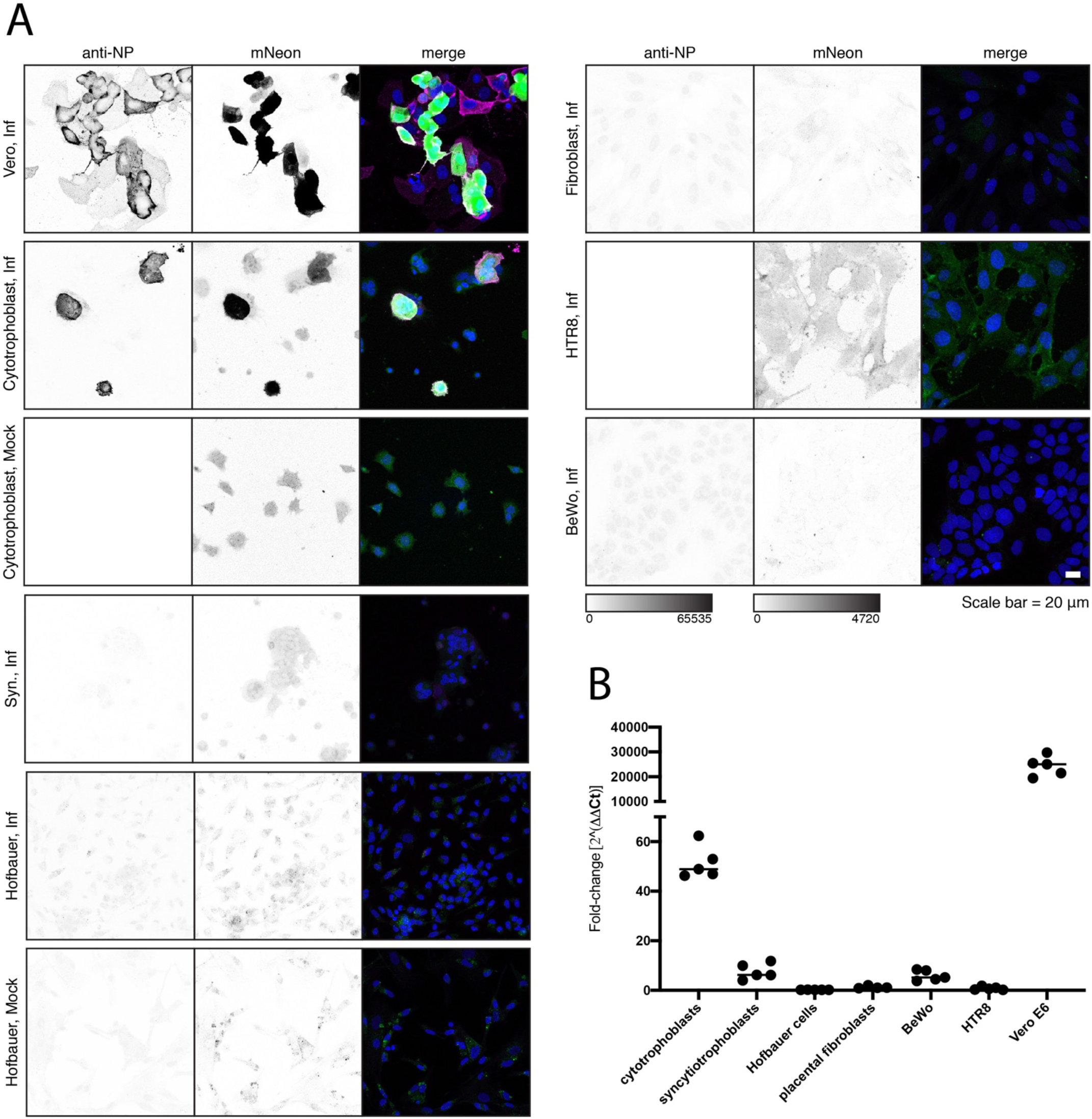
SARS-CoV-2 infection of placental cells in vitro. **(A)** Representative images of icSARS-CoV-2-mNG infection of primary placental cells, immortalized placental cell lines, and Vero E6 cells as measured by mNeonGreen expression and immunofluorescence staining of SARS-CoV-2 nucleocapsid (NP). Images are displayed as maximum intensity projections of z-stacks and grayscale bars indicate measured fluorescence intensity in arbitrary digital units. **(B)** Fold-change quantification of SARS-CoV-2 N1 by RT-qPCR at 24 hours post-infection. Cells were infected at an MOI of 5 for one hour and washed three times with PBS before the addition of fresh media. Cells were washed and collected at 24 hours post-infection. Data presented are representative results from one of three replicates.

Immortalized cell lines are commonly used as a model for placental cell types. The BeWo line, a human choriocarcinoma line, is used to model villous cytotrophoblasts. The HTR-8/SVneo line is derived from invasive extravillous cytotrophoblasts isolated from first trimester placenta and contains two cell populations(19, 20). Neither of these immortalized cell lines, BeWo and HTR-8/SVneo, exhibited significant infection with icSARS-CoV-2-mNG at 24 hours (Figure 3A).

While primary syncytiotrophoblasts and BeWo cells were not readily infected by icSARS-CoV-2-mNG *in vitro*, in rare cases, an individual cell exhibiting viral mNeonGreen fluorescence and NP staining (estimated to be <0.0001%) could be detected. These extremely rare positive cells were notably isolated cells that, in the case of the syncytiotrophoblast population, appear to be cytotrophoblast cells defective for syncytialization (Supplementary Figure 3). SARS-CoV-2 infection was thus not observed in any syncytialized placental cells in vitro. These results indicate that at least in the *ex vivo* context, cytotrophoblasts, but not other placental cell types, are infected by SARS-CoV-2.

### Transcriptional changes at the placenta during maternal COVID-19 reflect a localized inflammatory response to systemic SARS-CoV-2 infection

Despite the fact that cytotrophoblasts are permissive to SARS-CoV-2 infection *in vitro*, and that SARS-COV-2 infection of syncytiotrophoblasts has been demonstrated in isolated cases(10, 16, 21, 22), SARS-CoV-2 infection was not detected in the majority of the placentas tested in our case series. This was true even in women with high nasopharyngeal viral loads and with symptomatic SARS-CoV-2 infection, including those with severe complications. This absence of placental infection suggested the presence of a localized and effective anti-viral response in the placenta. We thus used bulk-RNA sequencing of placental villi to examine differences in placental gene expression between pregnant women with COVID-19 (n=5) and uninfected controls matched for maternal age, gestational age, maternal comorbidities, and mode of delivery (n=3).

In our comparison of the placental transcriptome in COVID-19 cases to matched controls, COVID-19 cases showed increased expression of genes associated with immune responses, suggesting a robust local response to respiratory infection, even in the absence of localized placental infection (p<0.05, Figure 4A). These changes in gene expression were largely shared among COVID-19 cases when compared to controls, as indicated by their grouping upon hierarchical clustering; placental transcriptomes from COVID-19 cases largely clustered together, and separately from healthy controls, with the exception of one case (COVID-1) (Figure 4A). Further analysis of differentially expressed genes by gene ontology indicated an enrichment in defense and immune response categories in COVID-19 cases compared to healthy controls (Figure 4B). The most significantly upregulated gene in the placenta during maternal SARS-CoV-2 infection was *HSPA1A*, which encodes the heat shock protein Hsp70 (Figure 4C), a proposed alarmin that has been previously implicated in placental vascular diseases and preeclampsia(23-25).

**Figure 4.**
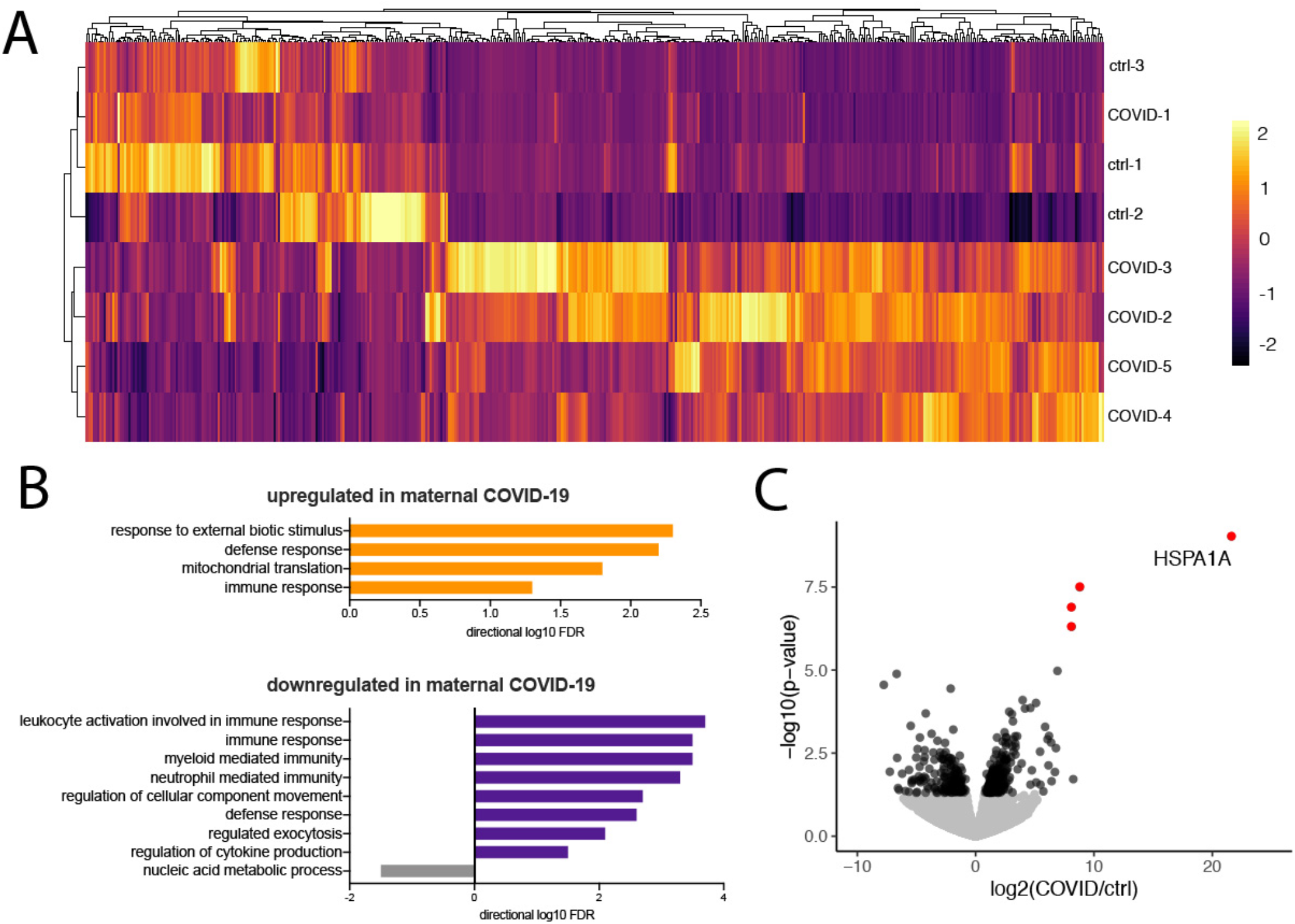
HSPA1A is significantly upregulated in maternal COVID. **(A)** Hierarchal clustering and heatmap of differentially expressed genes (p<0.05). Bulk RNA-seq was performed on placental villi isolated from control and maternal COVID cases. **(B)** Gene ontology of differentially expressed genes (p<0.05) in bulk RNA-seq. **(C)** Volcano plot indicating differentially expressed genes between control and maternal COVID groups from bulk RNA-seq. Significant hits are depicted in red (p_adj_<0.05) and black (p<0.05). Non-significant genes are shown in gray.

### Single cell transcriptomic profiling of the placenta reveals cell-type specific immune response to maternal SARS-CoV-2 infection

We next assessed COVID-19 associated transcriptomic changes in the placenta in a cell-type specific manner. To do so, we characterized placenta cells from hospitalized maternal acute COVID-19 cases (n=2) and matched controls (n=3) through unbiased, single cell RNA sequencing. A total of 83,378 cells were included in the analysis, 44,140 from placental villi and 39,238 from decidua parietalis. Placentas from these COVID-19 cases tested negative for SARS-CoV-2 by RT-qPCR. To further test for the presence of intracellular virus, open reading frames of SARS-CoV-2 (spike, ORF3a, envelope, membrane glycoprotein, ORF6, ORF7a, ORF8, nucleocapsid and ORF10) were added to the reference genome before alignment with CellRanger. No viral transcripts were detected in any of the placenta cells. We next performed unsupervised cluster analysis and represented single cell transcriptome data from COVID-19 cases and controls in UMAP space (Figure 5A).

**Figure 5.**
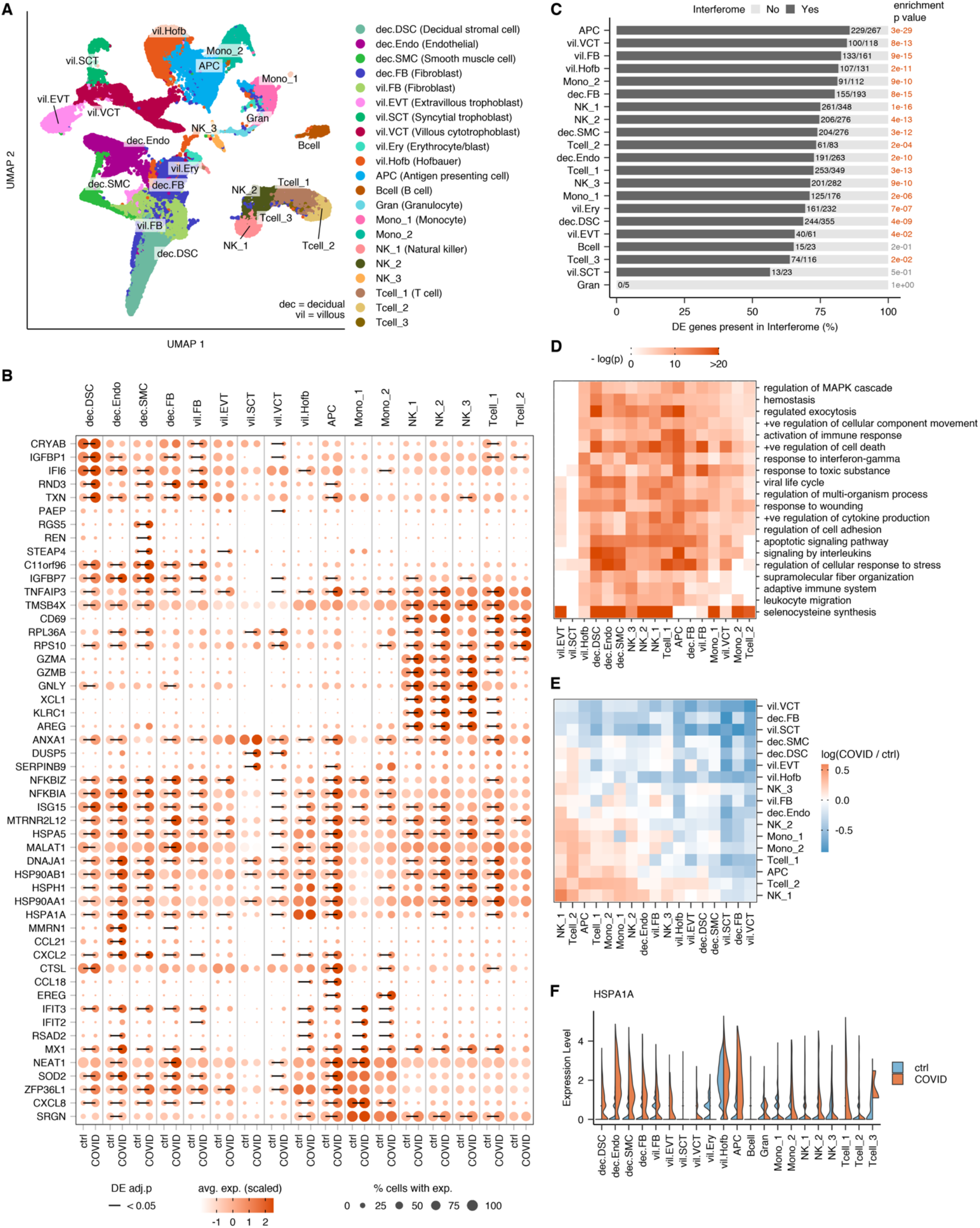
**(A)** UMAP projection of 83378 single placenta cells from COVID-19 cases (n=2 decidual and n=2 villous samples) and uninfected controls (n= 2 decidual and n = 3 villous samples). Cell type annotations based on correlation with reference datasets(26-28) followed by manual examination of marker genes. **(B)** Dotplot of the top 5 genes that are upregulated between COVID-19 and uninfected control samples for each annotated cell type based on fold-difference. Size of dots represents percent of cells in cluster expressing gene of interest; intensity of color reflects average scaled expression. Significantly altered expression between COVID-19 cases and controls (Bonferroni-adjusted, two-tailed, Wilcoxon rank-sum test *P* < 0.05) is marked by a solid black line. **(C)** Interferome analysis demonstrating the fraction of differentially expressed genes in each cell type that are interferon-responsive, in COVID-19 cases compared to controls; with *p* values for enrichment (observed over expected fraction) calculated using hypergeometric distribution. **(D)** Clustered heatmap showing the top enriched functional terms according to Metascape(76) among differentially expressed genes between COVID-19 and control samples in the annotated placental cell types. bars are colored to encode *p*-values of increasing statistical significance. **(E)** Heat map depicting the log-transformed ratio (COVID-19 cases over controls) of number of ligand-receptor interactions between all placental cell type pairs, inferred using the CellphoneDB repository of ligands, receptors and their interactions(33). Red indicates cell type pairs with more interactions in COVID-19 cases compared to control; blue indicates the opposite. **(F)** Violin plots of *HSPA1A* expression at the placental villi and maternal decidua obtained by scRNA-seq.

The classification of placental cells by single cell RNA sequencing is challenging because the placenta is comprised of multiple cell types, of both maternal and fetal origin, in highly active transcriptional states. We overcame this challenge by creating a reference set of cell type-specific transcriptomes for cell types present at the human fetal-maternal interface, using cell type-averaged expression values from three previously published placenta single cell RNA sequencing studies(26-28). We then compared single cell transcriptomes in our dataset to this composite reference dataset, and, after annotating clusters by comparison with the reference dataset, we manually examined clusters for marker gene expression and annotated remaining clusters. Through this approach we identified 21 distinct cell types in the placenta (Figure 5A).

Differential gene expression analysis revealed significantly altered gene expression in both immune and non-immune cell types in the placenta from COVID-19 cases (Figure 5B), including markedly increased expression of pro-inflammatory genes and chemokines. In placental NK cells during COVID-19, we found significant enrichment of genes encoding cytotoxic proteins, including *GZMA, GZMB*, and *GNLY*, as well as a tissue-repair growth factor, *AREG*. T cell subsets from COVID-19 cases upregulated *CD69*, a classical activation marker, as well as genes encoding ribosomal proteins, *RPL36A* and *RPS10*. Among endothelial cells, which have previously been implicated in COVID-19 pathogenesis, including COVID-19 associated thrombosis and vasculopathy(29), we found evidence for increased innate immune responses in COVID-19 cases compared to controls, including significant upregulation of *ISG15*, an interferon-induced protein that has been implicated as a central player in host antiviral responses, and *NFKBIA* and *NFKBIZ*, critical regulators of the NF-KB pathway. Notably, while we do not find significant expression of *ACE2* or *TMPRSS2* in placental cells in either COVID-19 cases or healthy controls, we do find widespread expression of *CTSL*, an alternative SARS-CoV-2 entry co-receptor, including in placental immune cells, fibroblasts, and trophoblasts, and find that *CTSL* expression is increased in decidual stromal cells and decidual antigen presenting cells in COVID-19.

Given the increasing evidence that hospitalized patients with COVID-19 demonstrate strong type-1 interferon responses(30, 31), we used Interferome(32), a database of interferon-regulated genes, to determine whether an interferon-driven inflammatory signature is displayed by the placenta. We find that placental cellular subsets demonstrate significantly increased expression of interferon-related genes in COVID-19 compared to healthy controls (Figure 5C). Pathway analysis of all differentially expressed genes likewise shows increased engagement of immune-related pathways in placental subsets from COVID-19 cases, as well as increases in synthesis of selenocysteine associated with the anti-oxidative response to oxidative stress (Figure 5D). Finally, we asked whether the transcriptional changes observed in placental tissue suggested altered cellular interactions in the placenta during COVID-19 compared to healthy conditions. Using CellphoneDB signaling network analysis(33), we found a significant increase in the number of interactions between immune cells at the maternal-fetal interface in COVID-19 cases when compared to controls. Among the strongest enriched relationships identified in COVID-19 cases were the interactions of T cells with monocytes and NK cells (Figure 5E), suggesting innate to adaptive immune cell communication in the local placental environment during maternal COVID-19.

Consistent with the bulk RNA-seq data, analysis of single-cell data indicated significant upregulation of *HSPA1A*, the gene encoding heat shop protein 70, in select placenta cellular subsets in COVID-19, including decidual APCs, decidual endothelial cells, and extravillous trophoblasts (Figure 5F).

## Discussion

Immune responses in the placenta can be a double-edged sword. These responses are critical for protecting the developing fetus from pathogen invasion, but, at the same time, placental inflammation itself may lead to pathological changes detrimental to pregnancy and fetal development(34-37). In this study, we demonstrate that a placental cell type is susceptible to SARS-CoV-2 infection *ex vivo*; that SARS-CoV-2 infection of the placenta *in vivo* is rare; and that a robust maternal immune response is mounted at the placenta even in the absence of placental infection.

While previous studies analyzing transcriptomic data have yielded mixed results regarding *ACE2* expression at the placenta(11-15), our immunohistochemical analysis conclusively demonstrated that ACE2 protein is present in the placenta despite low transcript levels. Furthermore, ACE2 protein expression is highest in the first trimester and decreases over the course of healthy pregnancy, indicating potential vulnerability to SARS-CoV-2 infection during early pregnancy. Surprisingly, we found that ACE2 expression appears to be widely expressed in the placenta of COVID-19 cases at term despite low levels of ACE2 in the placentas of healthy controls at term. The unique modifying factors that drive placental ACE2 expression during COVID-19 remain unknown; however, studies of ACE2 expression during other disease states, including COPD, suggest that ACE2 is upregulated under inflammatory conditions(38). Our data suggest that the hyperinflammatory state associated with COVID-19 may similarly increase ACE2 expression at the term placenta.

The presence of ACE2 at the syncytiotrophoblast layer of the placental villi is consistent with the finding that the *in vivo* distribution of SARS-CoV-2 in rare cases of placental infection is also observed predominantly at the syncytiotrophoblasts(10, 16, 21, 22). In contrast, we found that only cytotrophoblasts are infected by SARS-CoV-2 *in vitro*. Given that syncytiotrophoblasts originate from the spontaneous differentiation and fusion of cytotrophoblast stem cells(39), it is possible that removed from their *in vivo* context, these terminally differentiated cells are not capable of supporting a productive viral infection *in vitro*. Differences in susceptibility to viral infection *in vivo* versus *in vitro* have also been demonstrated for Zika virus(40). Notably, unlike Zika virus, and many other “TORCH” pathogens capable of causing congenital conditions following *in utero* exposure, SARS-CoV-2 does not appear to productively infect placental Hofbauer cells either *in vivo* or *in vitro*(41-43).

Despite the capacity of cytotrophoblasts to be infected *in vitro*, SARS-CoV-2 invasion of the placenta is only rarely observed *in vivo*. Indeed, only ∼13% of maternal COVID-19 cases we examined demonstrated placental infection with SARS-CoV-2. This may be due to previously reported low levels of SARS-CoV-2 viremia(44) (i.e., the absence of a direct route of infection to the placenta *in vivo*), or to variable ACE2 expression at term. Unfortunately, we were unable to screen for potential maternal SARS-CoV-2 viremia in our cohort. Nonetheless, even in the absence of placental infection, we observed localized gene expression differences in SARS-CoV-2-affected term placentas indicating a marked immune response to maternal respiratory infection distinctly manifesting at the maternal-fetal interface. The majority of these differentially expressed genes are interferon-regulated genes, demonstrating the capacity of the placenta to sense and respond to both local and distal infection.

We found that *HSPA1A* (Hsp70), is highly upregulated in the placenta during maternal COVID-19. Notably, Hsp70 has been proposed as a potential alarmin that has been shown *in vitro* to stimulate proinflammatory processes associated with parturition and pre-term birth(23, 45, 46). Hsp70 is associated with endothelial activation in placental vascular disease(25) and serum levels are increased in cases of preeclampsia(24, 47, 48). Hsp70 levels are furthermore significantly elevated in patients exhibiting hemolysis, elevated liver enzymes, and low platelet count (HELLP syndrome) compared to patients with severe preeclampsia without HELLP syndrome(24, 49). Intriguingly, there have been multiple reports of HELLP or HELLP-like syndrome in pregnant women affected by SARS-CoV-2 infection and COVID(10, 50, 51). While the interplay between COVID-19 and HELLP-like syndrome remains incompletely understood, these results suggest a potential common pathway for COVID-19 associated maternal morbidity and placental vascular diseases, including HELLP and preeclampsia. Moreover, extracellular Hsp70 is known to stimulate proinflammatory cytokines such as TNF-α, IL-1β, and IL-6(24).

We found increased intervillous fibrin deposition in the placenta in approximately one third of the COVID-19 cases. Intervillous fibrin is a pathological finding that increases with decreased maternal perfusion, increased maternal coagulability, and decreased thrombolytic function of the trophoblasts(52). Intervillous fibrin has been previously reported in cases of maternal COVID-19(10, 53), but the significance of this findings is unclear. One possibility may be that maternal SARS-CoV-2 infection activates the maternal endothelium, leading to a localized decrease in fibrinolysis, thereby causing fibrin buildup in a manner similar to that which is observed in preeclampsia (54). Alternatively, activation of immune cells in the placenta and circulating pro-inflammatory cytokines may trigger pro-coagulant signals in the local environment (55), inducing tissue factor synthesis from the syncytiotrophoblasts(56), ultimately leading to the accumulation of fibrin. Our single-cell transcriptomic analysis supports this hypothesis by demonstrating increased NK cell and endothelial cell expression of genes involved in supramolecular fiber organization pathways in placenta derived from COVID-19 cases.

This study is subject to several limitations. First, only 15 placentas were available for RT-qPCR analysis, and thus we had insufficient sample size to understand if specific clinical features (e.g., severe COVID-19 or duration of symptoms) correlated with the presence of local virus in the placenta. Furthermore, our analysis is limited in that we only assessed placenta from women who were infected with SARS-CoV-2 during the third trimester of pregnancy, and thus does not account for pathological and inflammatory changes at the placenta that result from infection during the first or second trimester. Indeed, our results demonstrating widespread ACE2 expression in the placenta during the first and second trimesters indicates that early pregnancy may be the most vulnerable time for SARS-CoV-2 induced placental pathology, and additional studies are needed to assess for potential placental and fetal abnormalities associated with infection during early pregnancy.

By characterizing changes at the maternal-fetal interface in the context of systemic infection, our research indicated that maternal SARS-CoV-2 infection at term is associated with an inflammatory state in the placenta that may contribute to poor pregnancy outcomes in COVID-19, even in the absence of viral invasion of the placenta(57). These immune responses in the placenta may serve to protect the placenta and fetus from infection, but they also have the potential to drive pathological changes with adverse consequences for developing embryos and fetuses, since *in utero* inflammation is associated with neurodevelopmental and cognitive disorders in affected offspring (36, 58). Mouse models of congenital viral infection have also shown that type I IFN signaling during early embryonic development can cause fetal demise(59), through the upregulation of IFITM proteins that interfere with cytotrophoblast fusion(60, 61). Further studies are therefore needed to assess the long-term consequences of SARS-CoV-2 associated immune activation in pregnant women regardless of local infectious status of the placenta.

## Methods

### Study participants

We searched the electronic medical record of Yale-New Haven Hospital for all women who tested positive for SARS-CoV-2 in the antepartum or peripartum period and presented for delivery hospitalization during the study period (March 27, 2020 to June 1, 2020). Women who were admitted to Yale New Haven Hospital Labor and Birth during the study period and who were positive for SARS-CoV-2 by nasopharyngeal swab RT-qPCR in the antepartum period or at the time of delivery hospitalization were approached for consent to donate additional tissue for research studies through the Yale IMPACT study (Implementing Medical and Public Health Action Against Coronavirus in CT). These participants provided informed consent, including for research studies of donated placental tissue. SARS-CoV-2 uninfected women (as determined by negative RT-qPCR testing of nasopharyngeal swab) were recruited during the study period and provided informed consent to donate placenta tissue to serve as uninfected controls for transcriptomic studies. Pre-pandemic histological controls were selected from pathology files at Yale New Haven Hospital and matched to the COVID-19 placental cases by maternal age, gestational age, and maternal co-morbidities. The study was approved by the Yale Institutional Review Board (protocol #2000027690 and 2000028550).

### SARS-CoV-2 S1 spike protein IgM and IgG serology testing

ELISA assays for IgG and IgM antibodies towards SARS-CoV-2 were performed on plasma as described by Amanat et al (62). Screening of 367 total plasma samples from all SARS-CoV-2-positive patients enrolled was performed using a 1:50 dilution.

### SARS-CoV-2 detection in placenta by RT-qPCR

Placenta samples were homogenized and centrifuged before nucleic acid was extracted using the MagMax Viral/Pathogen Nucleic Acid Isolation kit. SARS-CoV-2 was detected using a modified RT-qPCR assay with the N1, N2, and human RNase P (RP) primer-probe sets developed by the CDC and the NEB Luna Universal Probe One-Step RT-qPCR kit on the Bio-Rad CFX96 Touch Real-Time PCR Detection System(63). Each sample was extracted and tested in duplicate to confirm results. Placenta samples were considered positive by RT-qPCR if cycle threshold (CT) values for N1 and N2 were both <38, and with any value of RP. Samples were considered negative if N1 and N2 >38, and RP <38. Samples were considered invalid if N1 and N2 >38 and RP >38.

### Histopathological analysis of placenta

Pathology files at Yale New Haven Hospital were searched for placentas corresponding to cases of maternal COVID-19. All available placental cases were included in the histological study. Additionally, pathology files were searched for placentas from mothers without SARS-CoV-2 infection (pre-June 2019) to serve as historical controls. These controls were matched to study cases for maternal age, gestational age and maternal comorbidities. A total of 27 cases and 10 matched controls were assessed histologically. Of those, one case was of a dichorionic diamniotic twin pregnancy. The placentas of this twin pregnancy demonstrated extremely similar microscopic features and identical pathologic scores; thus, they were together considered as a single case in the statistical analysis. All placentas received with a requisition form for pathologic evaluation were immediately fixed in 10% neutral buffered formalin for three days. A total of six sections inclusive of at least two full thickness sections, peripheral membranes and umbilical cord were submitted for histologic examination.

Two pathologists (LI and RM) blinded to patient information and diagnostic report independently scored all H&E stained placental tissue for villitis (absent, low-grade or high-grade; if low-grade, focal or multifocal and if high-grade, patchy or diffuse), intervillositis (absent or present), increased intervillous fibrin (absent or present, defined as intervillous fibrin occupying >10% of a full thickness section on 20x low-power magnification), chorioamnionitis (absent or present; if present, maternal and fetal inflammatory responses were staged and graded), fetal and/or maternal vascular malperfusion (absent or present; if present, a histologic description was recorded) and increased decidual lymphocytes (absent or present, defined as clusters of >10 lymphocytes in >3 foci in the placental disc and/or peripheral membranes). Diagnostic criteria for villitis, chorioamnionitis, fetal vascular malperfusion (FVM) and maternal vascular malperfusion (MVM) followed those reported in the Amsterdam placental workshop group consensus statement(64). Cases with discrepant scoring results were reviewed and re-scored by both pathologists simultaneously. Results were correlated with maternal, fetal and placental COVID-19 status.

### Collection of placentas across gestation

First and second trimester specimens from the elective termination of pregnancy, ranging from 7 to 15 weeks of gestation (based on last menstrual period), were collected from otherwise healthy women with no known genetic or other abnormalities, as previously described(65). All women who provided first and second trimester placentas signed an informed consent (protocol #021-06-972) approved by the ethical committee of the Bnai Zion Medical Center, Haifa, Israel under Helsinki convention guidelines. Samples from normal term placentas were collected anonymously from healthy patients undergoing elective repeat Cesarean sections. All women who provided term placental samples signed an informed consent (Yale IRB protocol 1208010742). Specimens were fixed in 10% neutral buffered formalin (NBF) for at least one day and embedded in paraffin, after which 5 µm sections were placed on coated glass slides designed for immunohistochemistry (IHC) processing. Placentas of gestational age 18 to 23 weeks were obtained from non-genetic healthy terminations through the University of Pittsburgh Biospecimen Core (IRB#: PRO18010491).

### ACE2 Immunohistochemistry

The following rabbit polyclonal antibodies were used: anti-ACE2 (Abcam, Cambridge, MA, ab10 8252, used at 1 µg IgG/ml); and, as a negative control, normal rabbit serum (Sigma-Aldrich, St. Louis, MO, R9133, used at 1 µg IgG/ml). Antibody concentrations were chosen to produce strong staining in the positive cellular structures without background staining.

Slides were immunohistochemically stained as previously described(66) using MACH 2 Rabbit HRP-Polymer (Biocare Medical, RHRP520L, Pacheco, CA) to mark the sites of antibody binding with a brown deposit. To minimize run-to-run variability, replicate samples were stained simultaneously with one antibody. Positive control sections from a de-identified normal human kidney were analyzed concurrently with each batch of slides. The stained sections were counterstained with hematoxylin.

For quantification of immunoreactivity, slides were inspected microscopically using a raster pattern to ensure that the entire histologic section was examined. A modified histology score(67) was calculated by multiplying the percentage of trophoblasts that stained by the average staining intensity (ranging from 0-3), resulting in H-scores from 0 and 300.

### Preparation of decidua and placental villi for bulk and single-cell sequencing

Placentas were collected from Yale-New Haven Hospital and transferred to the laboratory for processing. Placental villi were isolated by sampling from midway between the chorionic and basal plates of the placenta. The decidua parietalis was isolated from the chorioamniotic membranes as previously described(68). The chorioamniotic membranes were dissected, rinsed in PBS, and placed with the chorion facing upward. Blood clots were removed using fine-point forceps and membranes were rinsed in PBS. A disposable cell scraper was used to gently remove the decidual layer from the membrane.

Dissected tissues were rinsed thoroughly in PBS and minced with scissors in a tissue digestion buffer of Liberase TM (0.28 WU/ml) and DNase I (30 μg/ml) in HBSS with Ca^2+^ and Mg^2+^. Finely minced tissue was enzymatically digested at 37°C for 1 hour with agitation, pipetting, and further mincing every 10 minutes until disaggregated. The suspension was passed through sterile gauze, centrifuged at 1000 x g for 5 minutes at 4°C to pellet cells, and washed with fresh digestion buffer. After centrifugation, the supernatant was aspirated and the cell pellet was resuspended in ACK lysing buffer for 5 minutes. Cells were centrifuged and resuspended in RPMI media before filtering through a 70-μm mesh cell strainer.

### Bulk RNA sequencing

Total RNA was prepared from freshly isolated placental villi as previously described(63). Depletion of rRNA, library preparation, and sequencing on the Illumina HiSeq 2500 platform were performed at the Yale Center for Genome Analysis (YCGA).

FASTQ files from HiSeq 2500 were analyzed using Kallisto v0.46.1(69) using the “-b 100 and -t 20” options to obtain transcript abundances in TPM and estimated counts. The kallisto index used during transcript quantification was built (31bp k-mer length) from the *Homo sapiens* transcriptome GRCh38 downloaded as a FASTA file from Ensembl (Ensembl.org). Transcripts were annotated using the Bioconductor package biomaRt v2.40.5(70) in R v3.6.2.

Read counts for individual transcripts were summarized for gene-level analysis using the Bioconductor package tximport with default parameters(71). Differential expression analysis was performed using DESeq2 with default parameters(72), comparing all 3rd trimester samples by maternal status.

### Single-cell RNA sequencing

#### Library preparation, data-preprocessing, and clustering

scRNA-seq libraries were generated using the 10x Chromium Single Cell 3’ Reagent Kit and libraries were sequenced using the Illumina NovaSeq platform. Sequencing results were demultiplexed into FASTQ files using the Cell Ranger (10x Genomics, 3.0.2) mkfastq function. Samples were aligned to GRCh38 10x genome and the count matrix was generated using the count function with default settings. Low quality cells containing <500 or >6000 genes detected were removed, as well cells with >20% of transcripts mapping to mitochondrial genes. Genes that were present in less than 3 cells were excluded from analysis. Gene expression values were then normalized, log-transformed, and scaled. Single cell transcriptomes from COVID-19 cases and healthy controls were pooled prior to unsupervised cluster analysis. Seurat version 3.1.5 with R version 3.4.2 was used for normalization, dimensionality reduction, clustering, and UMAP visualization, while Seurat version 3.2.2(73) with R version 4.0.2(74) was used for all downstream analyses.

#### Cluster annotation

Preliminary annotation of all clusters was performed by the similarity of their gene expression to annotated cell types in published single cell RNA-seq datasets of human fetal-maternal interface(26-28). For each cluster, mean of SCTransform-normalized(75) gene expression across cells was used as the cluster’s average gene expression. Spearman correlation coefficients between the cluster’s average expression and averaged expression of annotated cell types from all three reference datasets were calculated, and top three cell types with highest correlation coefficient were assigned as preliminary annotations of the clusters (Supp. Fig. 4). These annotations were further refined through manual examination of cluster marker genes (identified using Seurat function FindAllMarkers with options only.pos = TRUE and logfc.threshold = 0.25). Clusters that are highly similar to each other and have the same final cell type annotation, were then merged, resulting in the final set of 21 annotated clusters.

#### Differential gene expression

For each annotated cluster, differentially expressed genes between control and COVID-19 samples were identified using Wilcoxon rank-sum test as implemented in the Seurat function FindMarkers. Genes that met the following criteria were considered differentially expressed: absolute log-fold difference of 0.4 (corresponding to at least 50% difference in expression) and Bonferroni-adjusted two-tailed *p*-value less than 0.05.

#### Functional enrichment analyses

Two types of functional gene set enrichment analyses were performed, Interferome(32) and Metascape(76). For Interferome analysis, lists of differentially expressed genes in each cluster were searched against Interferome (version 2.01) with the default parameters except that the search was limited to human genes. Enrichment score for each cluster was calculated as the ratio between the observed fraction of differentially expressed genes found in Interferome database and the expected fraction, where the expected fraction is the ratio of total number of genes in the Interferome database over the total number of genes used in differential expression analysis. Enrichment *p*-values were calculated from the hypergeometric distribution using the R function phyper. Functional enrichment with Metascape was performed on lists of differentially expressed genes using the web application.

#### Ligand-receptor interactions

Ligand-receptor interactions were inferred using CellPhoneDB(33) separately for cells from control and COVID-19 samples. CellPhoneDB version 2.1.4 was used with Python version 3.6. For computational efficiency both control and COVID-19 data were subsampled to 5000 cells each.

### Placental explant preparation

Healthy term placentas were collected from scheduled cesarean sections performed at Yale-New Haven Hospital. Standard clinical criteria were used to exclude cases of infection and inflammation. 20mg of placental villous tissue was processed and rinsed in PBS. Explants were plated into a 24-well plate with 0.4-μm permeable transwell cell culture inserts (Corning) and maintained in F12:DMEM with 10% fetal bovine serum.

### Primary cell isolations from placenta

Isolation of Hofbauer cells and cytotrophoblasts from healthy term placentas was performed as previously described(77). Placentas from uncomplicated term pregnancies were brought to the laboratory within 30 minutes following elective cesarean section without labor at Yale-New Haven Hospital and processed immediately. Villous tissue was dissected, minced, and rinsed with PBS. Minced tissue was subjected to sequential enzymatic digestions in a solution of 0.25% trypsin and 0.2% DNase I at 37°C. Undigested tissue was removed by passage through gauze and a 100-μm sieve. Cells were resuspended in DMEM:F12 media with 10% FBS and 1% antibiotic/antimycotic.

Cytotrophoblasts were separated on a discontinuous gradient of Percoll (50%/45%/35%/30%) by centrifugation at 1000 x g for 20 minutes at room temperature. Cells migrating to the 35%/45% Percoll interface were recovered by centrifugation at 300 x g for 10 minutes at room temperature and immunopurified by negative selection using mouse anti-human CD9 antibody and mouse anti-human CD45 antibody. Following incubation with goat anti-mouse IgG-conjugated Dynabeads, contaminating cells were removed by exposure to a magnet.

Hofbauer cells were isolated from further digestion of trypsin-treated tissue with collagenase A and DNase I. Cells were pelleted, resuspended in RPMI, and loaded onto a discontinuous Percoll gradient (40%/35%/30%/20%) and centrifuged for 30 minutes. Cells from the 20%/25% to 30%/35% interfaces were combined and immunopurified by negative selection using sequential treatment with anti-EGFR and anti-CD10 antibodies conjugated to magnetic beads. Cells from the supernatant were plated and after 1 hour of incubation, floating and weakly adherent cells were removed.

Fibroblasts were obtained from cells removed during negative immunoselection of cytotrophoblasts and Hofbauer cells. Bead-cell mixtures were washed and cultured in media until confluency was reached. Following trypsinization of first passage cells, magnetic beads with attached cells (∼10% of population) were removed with a magnet. Passage 3 fibroblasts were used for experiments.

### SARS-CoV-2 infection

Primary placental cells were with icSARS-CoV-2 mNG(18) infected at an MOI of 5 for one hour at 37°C. Following infection, cells were washed three times in PBS before adding fresh media. Primary placental cells were maintained in F12:DMEM with 10% FBS supplemented with antibiotic/antimycotic. BeWo cells were maintained in F12K Kaighn’s modified media, HTR8 cells in RPMI media, and Vero E6 cells in F12:DMEM media, all with 10% FBS and antibiotic/antimycotic.

### Immunohistochemistry

Following dissection, placental tissue was rinsed in PBS and fixed in 10% neutral buffered formalin for 48-72 hours. Tissues were paraffin-embedded and cut into 5 μm thickness sections by Yale Pathology Tissue Services. Paraffin sections were heated for 30 minutes at 60°C and treated with xylenes followed by rehydration in decreasing concentrations of ethanol (100%, 90%, 80%, 70%). Antigen retrieval was performed by boiling in sodium citrate (pH 6.0) for 15 minutes and peroxidase activity was blocked with hydrogen peroxide for 10 minutes. Blocking was performed in 2.5% normal horse serum (Vector Laboratories) and incubated in primary antibody overnight at 4°C. Mouse anti-SARS-CoV-2 spike antibody (clone 1A9, GeneTex GTX632604) was used at a dilution of 1:400 and rabbit anti-ACE2 (clone EPR4435(2), Abcam ab108252) was used at 1:200 dilution. Secondary antibody and detection reagents from the VECTASTAIN Elite ABC-HRP Kit (Vector Laboratories PK-7200) were used according to manufacturer instructions. Sections were counterstained with Hematoxylin QS (Vector Laboratories H-3404), dehydrated in increasing concentrations of ethanol, cleared with xylenes, and mounted with VectaMount permanent mounting medium (Vector Laboratories H-5000).

### Immunofluorescence microscopy sample preparation and imaging

Placental cells on coverslips were fixed in fixed in 4% paraformaldehyde for 24 hours. Coverslips were blocked and permeabilized in 3% BSA (Sigma) and 0.1 % Triton X-100 (American Bioanalytical). Each sample was incubated overnight with anti-SARS-CoV-2-NP rabbit polyclonal antibody (GeneTex # GTX135357) at a dilution of 1:200. After washing in PBS, coverslips were incubated for 1 hour in a 1:500 dilution of Alexa 594 anti rabbit secondary antibody (Jackson ImmunoResearch 711-585-152), washed again in PBS and treated with 1 µg/mL Hoechst 33342 for 10 min and washed a final time in PBS. Samples were then mounted in DABCO/glycerol mounting media (Sigma) and imaged on a Leica Sp8 Laser Scanning Confocal Microscope equipped with a 40x N.A. 1.3 HC PL APO CS2 objective. Images are displayed as maximum intensity projections of z-stacks and a color bar is given in arbitrary digital units.

### SARS-CoV-2 S1 spike protein IgM and IgG serology testing

ELISA assays for IgG and IgM antibodies towards SARS-CoV-2 were performed on plasma as described by Amanat et al (62). Screening of plasma samples from all SARS-CoV-2-positive patients enrolled was performed using a 1:50 dilution.

### Statistical Analysis

One-way ANOVA was used to compare clinical and demographic features between the three groups presented in Table 1. Chi-square tests were used for statistical comparisons of histologic features of cases versus controls. Differences were claimed as statistically significant when the p value was less than 0.05.

## Data Availability

Raw sequence data will be deposited to the Gene Expression Omnibus (GEO) repository at the National Center for Biotechnology Information (NCBI).

## Data availability

Single cell RNA sequencing analysis code is available at https://github.com/archavan/covid-placenta

## Acknowledgments

We thank Vikki Abrahams for the contribution of placental cell lines (HTR8, courtesy of Charles Graham, and BeWo). We also thank Craig Wilen for supplying icSARS-CoV-2-mNG virus. Research support was provided by the Yale University Reproductive Sciences (YURS) Biobank, Yale Pathology Tissue Services (YPTS), and the Yale Center for Genome Analysis (YCGA). This project used the University of Pittsburgh Medical Center (UPMC) Hillman Cancer Center and Tissue and Research Pathology/Pitt Biospecimen Core shared resource which is supported in part by award P30CA047904.

ALC is supported by the National Institutes of Health (NIH) under Medical Scientist Training Program (MSTP) grant T32GM007205 and National Institute of Child Health & Development (NICHD) grant F30HD093350. ARC is supported by Irvington Postdoctoral Fellowship from Cancer Research Institute. AI is an investigator of the Howard Hughes Medical Institute. This work was supported by NIH K23MH118999 (SFF), R01AI157488 (SFF and AI), U01DA040588-05S (KMN), the Beatrice Kleinberg Neuwirth Fund (AK), and Fast Grant funding support from the Emergent Ventures at the Mercatus Center (AI). No NIH funding was used in the acquisition of placental tissue derived from healthy terminations.

Yale IMPACT Team Members: Charles Dela Cruz, Allison Nelson, Anne L. Wyllie, Melissa Campbell, Elizabeth B. White, Rebecca Earnest, Sarah Lapidus, Joseph Lim, Maura Nakahata, Angela Nunez, Denise Shepard, Irene Matos, Yvette Strong, Kelly Anastasio, Kristina Brower, Maxine Kuang, Camila Odio, Bertie Geng, Maksym Minasyan, Melissa Linehan, Anjelica Martin, Tyler Rice, William Khoury-Hanold, Jessica Nouws, David McDonald, Kadi-Ann Rose, Yiyun Cao, Lokesh Sharma, Mikhail Smolgovsky, Abeer Obaid, Giuseppe Deluliis, Hong-Jai Park, Nicole Sonnert, Sofia Velazquez, Xiaohua Peng, Michael H. Askenase, Codruta Todeasa, Mollyc L. Bucklin, Maria Batsu, Adam J. Moore, Eric Y. Wang.

## Author Contributions

ALC contributed to the design and implementation of the research, execution of experiments, analysis of the results, and writing of the manuscript. ARC analyzed the single cell RNA sequencing, and edited the manuscript. PV identified and consented cases and controls, collected biospecimens, and analyzed clinical and demographic information. LI and RM performed histological analyses. EC, HJL, and KMN contributed to in vitro studies. KM assisted with IHC experiments. ZT and SG performed placental cell isolations. SP and WJLC contributed to bulk RNA sequencing analyses. ES contributed to single cell RNA sequencing experiments and analysis. CBFV and NDG performed and analyzed RT-qPCR on placental tissue. FM and AR performed plasma antibody assays. KC, SB, JT, WS, JF, MCM, and LK assisted with case identification and biospecimen collection. ACM assisted with biospecimen collection and processing. AIK contributed to study design and edited the manuscript. HJK performed and analyzed IHC experiments, contributed to histological analyses and interpretation of results, and edited the manuscript. AI and SFF designed and supervised the study.

---

**Supplementary Table 1.** RT-qPCR results for placenta samples that tested positive by the modified CDC assay. Total nucleic acid was extracted from each sample and tested by RT-qPCR in duplicate. Reported are Ct values for the primer-probe sets N1, N2, and RP.

| Sample_ID | Sample_Type | Replicate | N1 | N2 | RP | Result |
| --- | --- | --- | --- | --- | --- | --- |
| #1 | Placenta | 1 | 13.9 | 15.3 | 21.3 | Positive |
|  | Umbilical Cord | 1 | 30.6 | 30.6 | 28.2 | Positive |
| #2 | Placenta Fetal | 1 | 31.6 | 33.9 | 17.4 | Positive |
|  |  | 2 | 29.3 | 31.2 | 17.2 | Positive |
|  | Placenta Maternal | 1 | 37.2 | 37.7 | 18.0 | Positive |
|  |  | 2 | ND | ND | 18.6 | Negative |
|  | Placenta Membrane | 1 | 33.1 | 33.8 | 19.4 | Positive |
|  |  | 2 | 32.0 | 32.6 | 18.8 | Positive |
|  | Umbilical Cord | 1 | 35.0 | 35.6 | 21.2 | Positive |
|  |  | 2 | 35.6 | 35.2 | 21.0 | Positive |

**Supplementary Table 2.** Histological features of placenta from COVID-19 cases and matched controls.

| **Microscopic Features** |  |  | **P-value** |
| --- | --- | --- | --- |
|  | **Cases**<br>**(n=27)** | **Controls**<br>**(n=10)** |  |
| **Increased Decidual Lymphocytes** | 7/27 | 2/10 | 0.71 |
| **Maternal Vascular Malperfusion** | 6/27 | 0/10 | 0.10 |
| **Fetal Vascular Malperfusion** | 3/27 | 0/10 | 0.27 |
| **Chorioamnionitis** | 5/27 | 0/10 | 0.14 |
| **Increased Intervillous Fibrin** | 9/27 | 0/10 | **0.04** |
| **Intervillositis** | 0/27 | 0/10 | NA |
| **Villitis** | 5/27 | 0/10 | 0.14 |

**Supplementary Figure 1.**
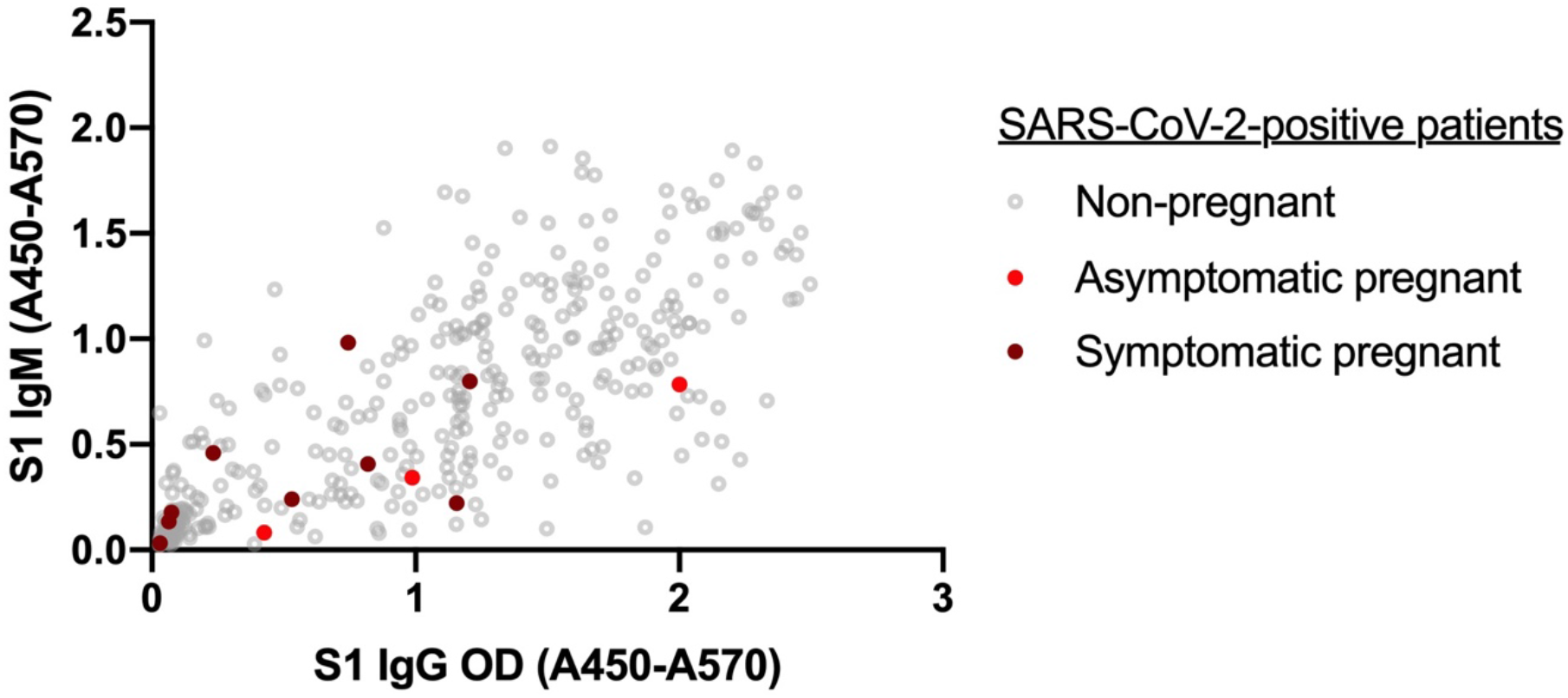
SARS-CoV-2 spike S1 protein-specific IgG and IgM (anti-S1-IgG and -IgM) ELISA absorbance values in 12 pregnant SARS-CoV-2-positive women compared to 355 non-pregnant SARS-CoV-2-positive individuals.

**Supplementary Figure 2.**
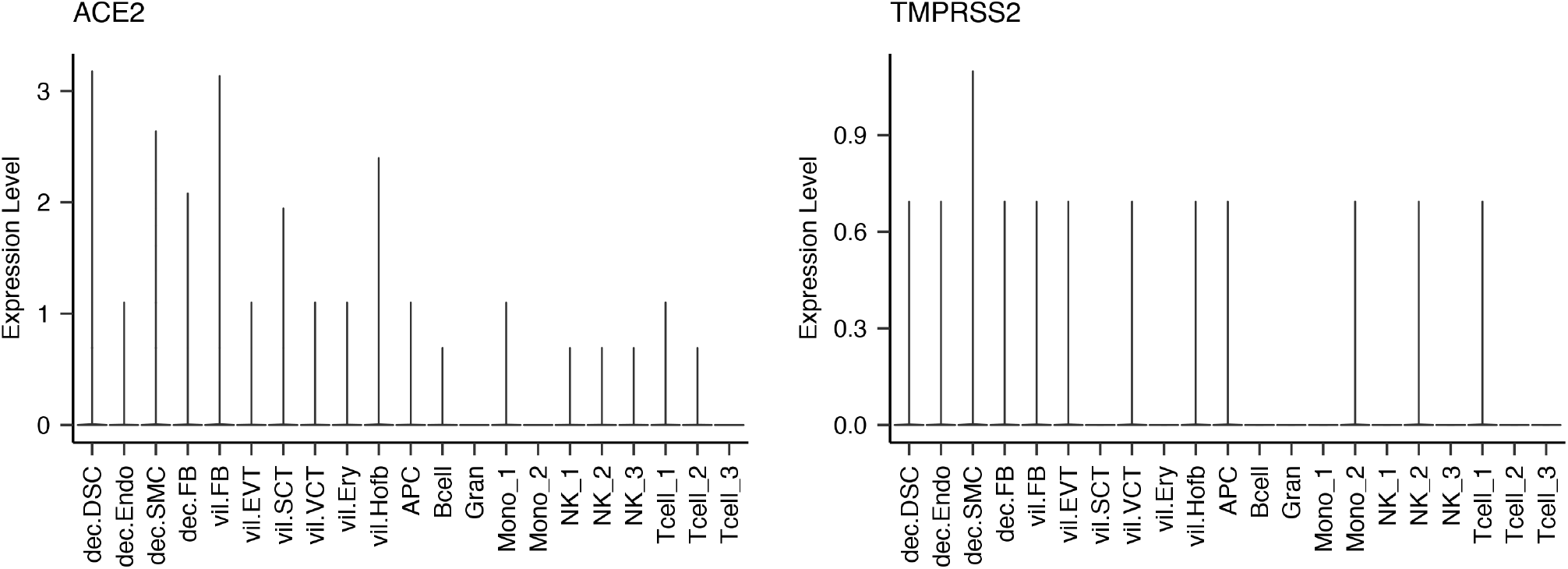
Violin plot of ACE2 and TMPRSS2 expression in placenta cell subsets.

**Supplementary Figure 3.**
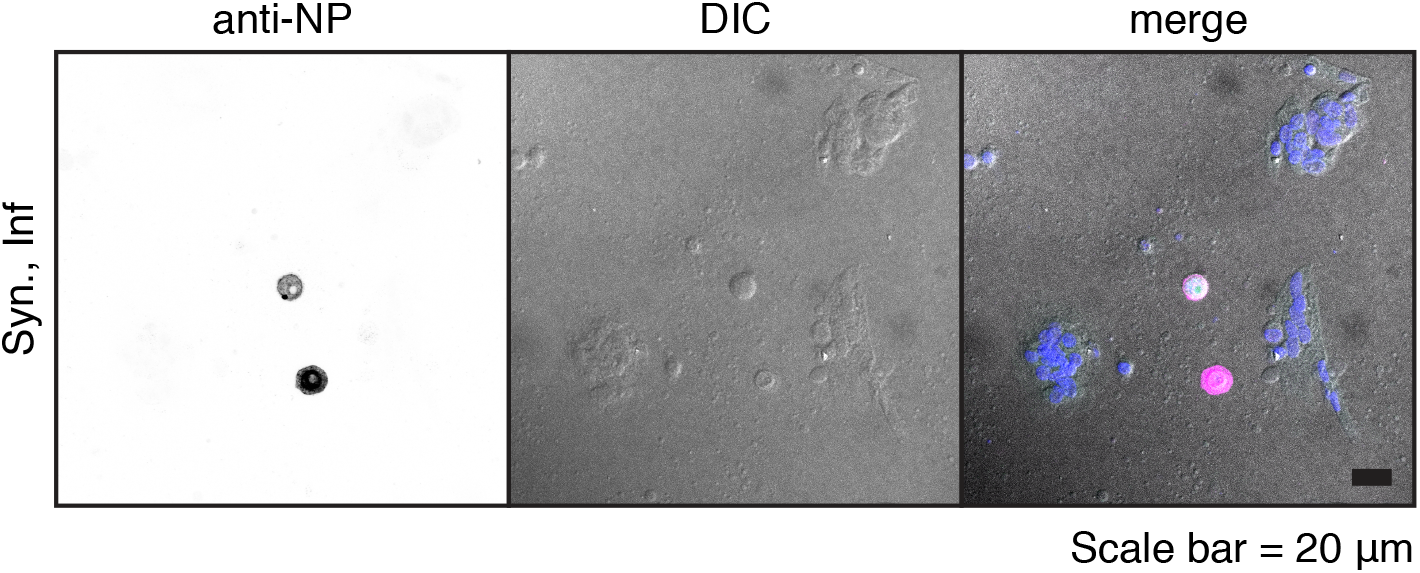
SARS-CoV-2 infection of primary syncytiotrophoblasts. Primary syncytiotrophoblasts were derived from primary isolated cytotrophoblasts allowed to spontaneously differentiate for 72-96 hours. Following the differentiation period, cells were infected at an MOI of 5 for one hour and washed three times with PBS before the addition of fresh media. Shown are two individual cells as detected by immunofluorescence following staining with anti-NP antibody (left panel). Differential interference contrast (DIC, center panel) image shows these two cells are excluded from the surrounding syncytia. Merged image (right panel) shows pseudocolor labeling of syncytialized cells in purple and infected NP-labeled individual cells in pink.

**Supplementary Figure 4.**
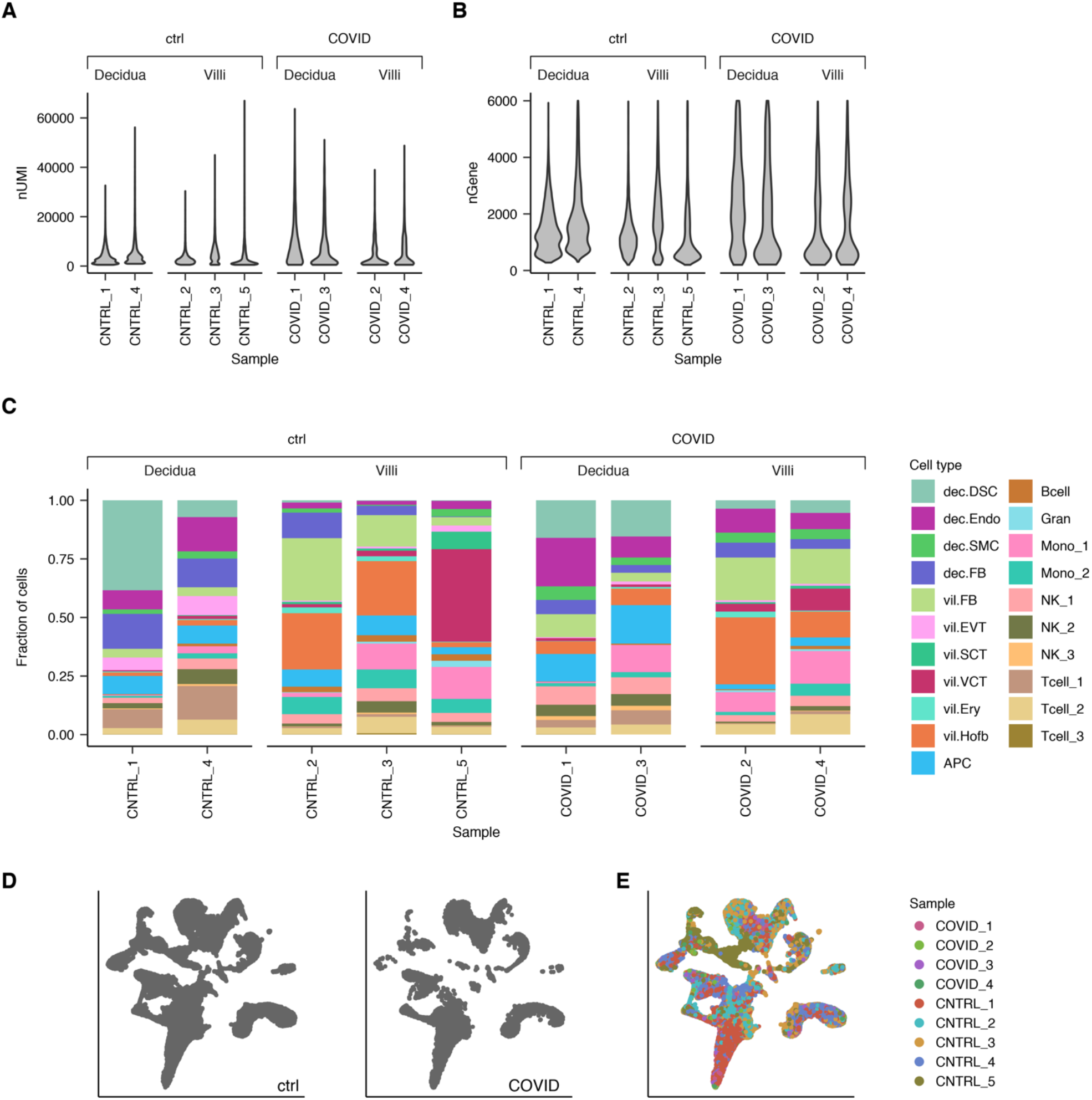
Quality control metrics for single cell RNA-seq samples. Violin plots showing the distributions of **(A)** the number of unique molecular identifier (UMI) counts per cell for all samples and **(B)** the number of detected genes per cell for all samples. **(C)** Stacked bar plots showing the cellular composition of each sample. Fraction of total cells made up by each annotated cell type is plotted on the Y axis. **(D)** UMAP projection of cells split by COVID-19 status showing that most annotated cell types are represented in both control and COVID-19 samples. **(E)** UMAP projection of cells colored by the sample of origin.

**Supplementary Figure 5.**
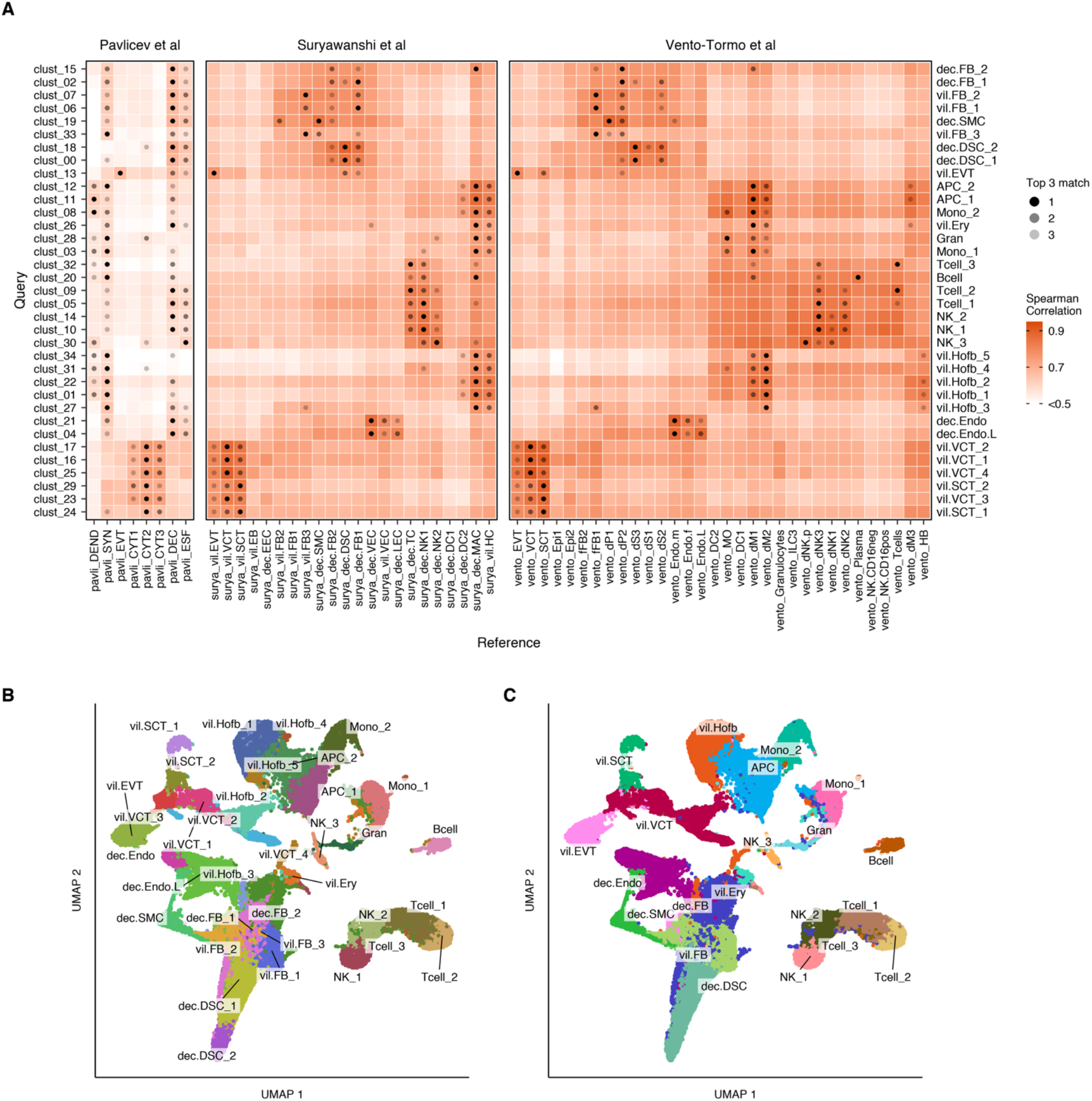
Annotation of single cell RNA-seq clusters based on correlation with reference datasets. **(A)** Heatmap showing spearman correlation coefficients between averaged gene expression of each cluster and that of annotated cell types from three single cell RNA-seq datasets^1–3^ used as references. For each query cluster, three annotated cell types with highest correlation from each reference dataset are marked as top three matches, represented by grey dots. Reference-based annotation of the clusters was refined with manual verification of marker gene expression. Refined annotations are shown on the right-hand side of the panel. **(B)** Annotation of all 35 clusters shown on UMAP projection. **(C)** Highly similar clusters were merged to get a final set of annotations resulting in 21 annotated clusters, presented on UMAP projection.

**Supplementary Figure 6.**
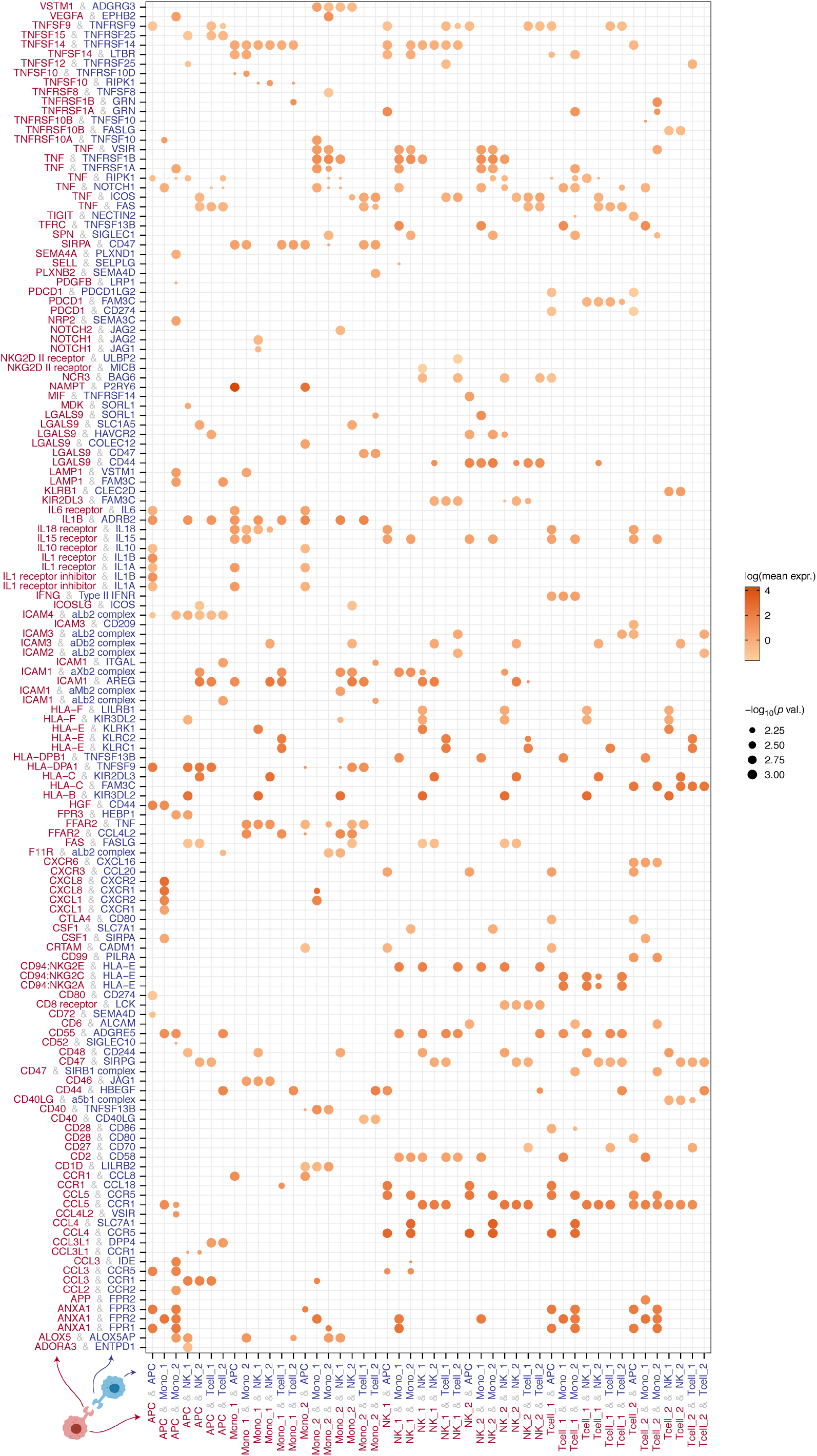
Dotplot showing ligand-receptor interactions, inferred using CellPhoneDB^4^, present in COVID-19 samples but absent in control samples. Pairs of cell types are on the X axis and pairs of ligands and receptors are on the Y axis, such that the first element of the latter is expressed in the first cell type (shown in red text) and the second element in the second cell type (shown in blue text). Intensity of the color of the dots represents mean of the expression of the ligand and the receptor in the respective cell types (mean is set to zero if either ligand or receptor has zero expression), and the size of the dot represents the *p*-value for the specificity of the interaction. Only the interactions with *p*-value < 0.01 are shown here.

## Notes

### Competing Interest Statement

The authors have declared no competing interest.

### Author Declarations

Informed consent was given and signed by all patients. All samples collected were approved by the respective Institutional Review Boards at Yale (protocol 1208010742), Bnai Zion Medical Center (protocol 021-06-972), and the University of Pittsburgh (PRO18010491).

## References

1. Zambrano LD, Ellington S, Strid P, Galang RR, Oduyebo T, Tong VT, et al. Update: Characteristics of Symptomatic Women of Reproductive Age with Laboratory-Confirmed SARS-CoV-2 Infection by Pregnancy Status - United States, January 22-October 3, 2020. MMWR Morb Mortal Wkly Rep. 2020;69(44):1641–7.

2. Ahlberg M, Neovius M, Saltvedt S, Soderling J, Pettersson K, Brandkvist C, et al. Association of SARS-CoV-2 Test Status and Pregnancy Outcomes. JAMA. 2020.

3. Abasse S, Essabar L, Costin T, Mahisatra V, Kaci M, Braconnier A, et al. Neonatal COVID-19 Pneumonia: Report of the First Case in a Preterm Neonate in Mayotte, an Overseas Department of France. Children (Basel). 2020;7(8).

4. Zimmermann P, and Curtis N. COVID-19 in Children, Pregnancy and Neonates: A Review of Epidemiologic and Clinical Features. Pediatr Infect Dis J. 2020;39(6):469–77.

5. Barthold SW, Beck DS, and Smith AL. Mouse hepatitis virus and host determinants of vertical transmission and maternally-derived passive immunity in mice. Arch Virol. 1988;100(3-4):171–83.

6. Cardenas I, Means RE, Aldo P, Koga K, Lang SM, Booth CJ, et al. Viral infection of the placenta leads to fetal inflammation and sensitization to bacterial products predisposing to preterm labor. J Immunol. 2010;185(2):1248–57.

7. Ng WF, Wong SF, Lam A, Mak YF, Yao H, Lee KC, et al. The placentas of patients with severe acute respiratory syndrome: a pathophysiological evaluation. Pathology. 2006;38(3):210–8.

8. Baud D, Greub G, Favre G, Gengler C, Jaton K, Dubruc E, et al. Second-Trimester Miscarriage in a Pregnant Woman With SARS-CoV-2 Infection. JAMA. 2020;323(21):2198–200.

9. Hecht JL, Quade B, Deshpande V, Mino-Kenudson M, Ting DT, Desai N, et al. SARS-CoV-2 can infect the placenta and is not associated with specific placental histopathology: a series of 19 placentas from COVID-19-positive mothers. Mod Pathol. 2020;33(11):2092–103.

10. Hosier H, Farhadian SF, Morotti RA, Deshmukh U, Lu-Culligan A, Campbell KH, et al. SARS-CoV-2 infection of the placenta. J Clin Invest. 2020;130(9):4947–53.

11. Pique-Regi R, Romero R, Tarca AL, Luca F, Xu Y, Alazizi A, et al. Does the human placenta express the canonical cell entry mediators for SARS-CoV-2? Elife. 2020;9.

12. Li M, Chen L, Zhang J, Xiong C, and Li X. The SARS-CoV-2 receptor ACE2 expression of maternal-fetal interface and fetal organs by single-cell transcriptome study. PLoS One. 2020;15(4):e0230295.

13. Ashary N, Bhide A, Chakraborty P, Colaco S, Mishra A, Chhabria K, et al. Single-Cell RNA-seq Identifies Cell Subsets in Human Placenta That Highly Expresses Factors Driving Pathogenesis of SARS-CoV-2. Front Cell Dev Biol. 2020;8:783.

14. Singh M, Bansal V, and Feschotte C. A Single-Cell RNA Expression Map of Human Coronavirus Entry Factors. Cell Rep. 2020;32(12):108175.

15. Hikmet F, Mear L, Edvinsson A, Micke P, Uhlen M, and Lindskog C. The protein expression profile of ACE2 in human tissues. Mol Syst Biol. 2020;16(7):e9610.

16. Patane L, Morotti D, Giunta MR, Sigismondi C, Piccoli MG, Frigerio L, et al. Vertical transmission of coronavirus disease 2019: severe acute respiratory syndrome coronavirus 2 RNA on the fetal side of the placenta in pregnancies with coronavirus disease 2019-positive mothers and neonates at birth. Am J Obstet Gynecol MFM. 2020;2(3):100145.

17. Vivanti AJ, Vauloup-Fellous C, Prevot S, Zupan V, Suffee C, Do Cao J, et al. Transplacental transmission of SARS-CoV-2 infection. Nat Commun. 2020;11(1):3572.

18. Xie X, Muruato A, Lokugamage KG, Narayanan K, Zhang X, Zou J, et al. An Infectious cDNA Clone of SARS-CoV-2. Cell Host Microbe. 2020;27(5):841–8 e3.

19. Graham CH, Hawley TS, Hawley RG, MacDougall JR, Kerbel RS, Khoo N, et al. Establishment and characterization of first trimester human trophoblast cells with extended lifespan. Exp Cell Res. 1993;206(2):204–11.

20. Abou-Kheir W, Barrak J, Hadadeh O, and Daoud G. HTR-8/SVneo cell line contains a mixed population of cells. Placenta. 2017;50:1–7.

21. Taglauer E, Benarroch Y, Rop K, Barnett E, Sabharwal V, Yarrington C, et al. Consistent localization of SARS-CoV-2 spike glycoprotein and ACE2 over TMPRSS2 predominance in placental villi of 15 COVID-19 positive maternal-fetal dyads. Placenta. 2020;100:69–74.

22. Facchetti F, Bugatti M, Drera E, Tripodo C, Sartori E, Cancila V, et al. SARS-CoV2 vertical transmission with adverse effects on the newborn revealed through integrated immunohistochemical, electron microscopy and molecular analyses of Placenta. EBioMedicine. 2020;59:102951.

23. Brien ME, Baker B, Duval C, Gaudreault V, Jones RL, and Girard S. Alarmins at the maternal-fetal interface: involvement of inflammation in placental dysfunction and pregnancy complications (1). Can J Physiol Pharmacol. 2019;97(3):206–12.

24. Molvarec A, Tamasi L, Losonczy G, Madach K, Prohaszka Z, and Rigo J Jr., Circulating heat shock protein 70 (HSPA1A) in normal and pathological pregnancies. Cell Stress Chaperones. 2010;15(3):237–47.

25. Liu Y, Li N, You L, Liu X, Li H, and Wang X. HSP70 is associated with endothelial activation in placental vascular diseases. Mol Med. 2008;14(9-10):561-6.

26. Vento-Tormo R, Efremova M, Botting RA, Turco MY, Vento-Tormo M, Meyer KB, et al. Single-cell reconstruction of the early maternal-fetal interface in humans. Nature. 2018;563(7731):347–53.

27. Pavlicev M, Wagner GP, Chavan AR, Owens K, Maziarz J, Dunn-Fletcher C, et al. Single-cell transcriptomics of the human placenta: inferring the cell communication network of the maternal-fetal interface. Genome Res. 2017;27(3):349–61.

28. Suryawanshi H, Morozov P, Straus A, Sahasrabudhe N, Max KEA, Garzia A, et al. A single-cell survey of the human first-trimester placenta and decidua. Sci Adv. 2018;4(10):eaau4788.

29. Goshua G, Pine AB, Meizlish ML, Chang CH, Zhang H, Bahel P, et al. Endotheliopathy in COVID-19-associated coagulopathy: evidence from a single-centre, cross-sectional study. Lancet Haematol. 2020;7(8):e575–e82.

30. Acharya D, Liu G, and Gack MU. Dysregulation of type I interferon responses in COVID-19. Nat Rev Immunol. 2020;20(7):397–8.

31. Lee JS, and Shin EC. The type I interferon response in COVID-19: implications for treatment. Nat Rev Immunol. 2020;20(10):585–6.

32. Rusinova I, Forster S, Yu S, Kannan A, Masse M, Cumming H, et al. Interferome v2.0: an updated database of annotated interferon-regulated genes. Nucleic Acids Res. 2013;41(Database issue):D1040–6.

33. Efremova M, Vento-Tormo M, Teichmann SA, and Vento-Tormo R. CellPhoneDB: inferring cell-cell communication from combined expression of multi-subunit ligand-receptor complexes. Nat Protoc. 2020;15(4):1484–506.

34. Nadeau-Vallee M, Obari D, Palacios J, Brien ME, Duval C, Chemtob S, et al. Sterile inflammation and pregnancy complications: a review. Reproduction. 2016;152(6):R277–R92.

35. Weckman AM, Ngai M, Wright J, McDonald CR, and Kain KC. The Impact of Infection in Pregnancy on Placental Vascular Development and Adverse Birth Outcomes. Front Microbiol. 2019;10:1924.

36. Lu-Culligan A, and Iwasaki A. The Role of Immune Factors in Shaping Fetal Neurodevelopment. Annu Rev Cell Dev Biol. 2020;36:441–68.

37. Yockey LJ, Lucas C, and Iwasaki A. Contributions of maternal and fetal antiviral immunity in congenital disease. Science. 2020;368(6491):608–12.

38. Leung JM, Yang CX, Tam A, Shaipanich T, Hackett TL, Singhera GK, et al. ACE-2 expression in the small airway epithelia of smokers and COPD patients: implications for COVID-19. Eur Respir J. 2020;55(5).

39. Kliman HJ, Nestler JE, Sermasi E, Sanger JM, and Strauss JF, 3rd. Purification, characterization, and in vitro differentiation of cytotrophoblasts from human term placentae. Endocrinology. 1986;118(4):1567–82.

40. Jurado KA, Simoni MK, Tang Z, Uraki R, Hwang J, Householder S, et al. Zika virus productively infects primary human placenta-specific macrophages. JCI Insight. 2016;1(13).

41. Reyes L, and Golos TG. Hofbauer Cells: Their Role in Healthy and Complicated Pregnancy. Front Immunol. 2018;9:2628.

42. Simoni MK, Jurado KA, Abrahams VM, Fikrig E, and Guller S. Zika virus infection of Hofbauer cells. Am J Reprod Immunol. 2017;77(2).

43. Coyne CB, and Lazear HM. Zika virus -reigniting the TORCH. Nat Rev Microbiol. 2016;14(11):707–15.

44. Kim JM, Kim HM, Lee EJ, Jo HJ, Yoon Y, Lee NJ, et al. Detection and Isolation of SARS-CoV-2 in Serum, Urine, and Stool Specimens of COVID-19 Patients from the Republic of Korea. Osong Public Health Res Perspect. 2020;11(3):112–7.

45. Pockley AG. Heat shock proteins as regulators of the immune response. The Lancet. 2003;362(9382):469–76.

46. Geng J, Li H, Huang C, Chai J, Zheng R, Li F, et al. Functional analysis of HSPA1A and HSPA8 in parturition. Biochem Biophys Res Commun. 2017;483(1):371–9.

47. Hromadnikova I, Dvorakova L, Kotlabova K, Kestlerova A, Hympanova L, Novotna V, et al. Circulating heat shock protein mRNA profile in gestational hypertension, pre-eclampsia & foetal growth restriction. Indian J Med Res. 2016;144(2):229–37.

48. Saghafi N, Pourali L, Ghavami Ghanbarabadi V, Mirzamarjani F, and Mirteimouri M. Serum heat shock protein 70 in preeclampsia and normal pregnancy: A systematic review and meta-analysis. Int J Reprod Biomed. 2018;16(1):1–8.

49. Molvarec A, Rigo J Jr.,, Nagy B, Walentin S, Szalay J, Fust G, et al. Serum heat shock protein 70 levels are decreased in normal human pregnancy. J Reprod Immunol. 2007;74(1-2):163–9.

50. Figueras F, LLurba E, Martinez-Portilla R, Mora J, Crispi F, and Gratacos E. COVID-19 causing HELLP-like syndrome in pregnancy and role of angiogenic factors for differential diagnosis. medRxiv. 2020:2020.07.10.20133801.

51. Futterman I, Toaff M, Navi L, and Clare CA. COVID-19 and HELLP: Overlapping Clinical Pictures in Two Gravid Patients. AJP Rep. 2020;10(2):e179–e82.

52. Fox H. Perivillous fibrin deposition in the human placenta. Am J Obstet Gynecol. 1967;98(2):245–51.

53. Shanes ED, Mithal LB, Otero S, Azad HA, Miller ES, and Goldstein JA. Placental Pathology in COVID-19. Am J Clin Pathol. 2020;154(1):23–32.

54. Stalker AL. Fibrin deposition in pregnancy. J Clin Pathol Suppl (R Coll Pathol). 1976;10:70–6.

55. Li M, and Huang SJ. Innate immunity, coagulation and placenta-related adverse pregnancy outcomes. Thromb Res. 2009;124(6):656–62.

56. Aharon A, Brenner B, Katz T, Miyagi Y, and Lanir N. Tissue factor and tissue factor pathway inhibitor levels in trophoblast cells: implications for placental hemostasis. Thromb Haemost. 2004;92(4):776–86.

57. Chaiworapongsa T, Erez O, Kusanovic JP, Vaisbuch E, Mazaki-Tovi S, Gotsch F, et al. Amniotic fluid heat shock protein 70 concentration in histologic chorioamnionitis, term and preterm parturition. J Matern Fetal Neonatal Med. 2008;21(7):449–61.

58. Makinson R, Lloyd K, Grissom N, and Reyes TM. Exposure to in utero inflammation increases locomotor activity, alters cognitive performance and drives vulnerability to cognitive performance deficits after acute immune activation. Brain Behav Immun. 2019;80:56–65.

59. Yockey LJ, Jurado KA, Arora N, Millet A, Rakib T, Milano KM, et al. Type I interferons instigate fetal demise after Zika virus infection. Sci Immunol. 2018;3(19).

60. Zani A, Zhang L, McMichael TM, Kenney AD, Chemudupati M, Kwiek JJ, et al. Interferon-induced transmembrane proteins inhibit cell fusion mediated by trophoblast syncytins. J Biol Chem. 2019;294(52):19844–51.

61. Buchrieser J, Degrelle SA, Couderc T, Nevers Q, Disson O, Manet C, et al. IFITM proteins inhibit placental syncytiotrophoblast formation and promote fetal demise. Science. 2019;365(6449):176–80.

62. Amanat F, Nguyen T, Chromikova V, Strohmeier S, Stadlbauer D, Javier A, et al. A serological assay to detect SARS-CoV-2 seroconversion in humans. medRxiv. 2020:2020.03.17.20037713.

63. Vogels CBF, Brito AF, Wyllie AL, Fauver JR, Ott IM, Kalinich CC, et al. Analytical sensitivity and efficiency comparisons of SARS-CoV-2 RT-qPCR primer-probe sets. Nat Microbiol. 2020;5(10):1299–305.

64. Khong TY, Mooney EE, Ariel I, Balmus NC, Boyd TK, Brundler MA, et al. Sampling and Definitions of Placental Lesions: Amsterdam Placental Workshop Group Consensus Statement. Arch Pathol Lab Med. 2016;140(7):698–713.

65. Kliman HJ, Sammar M, Grimpel YI, Lynch SK, Milano KM, Pick E, et al. Placental protein 13 and decidual zones of necrosis: an immunologic diversion that may be linked to preeclampsia. Reprod Sci. 2012;19(1):16–30.

66. Kliman HJ, Quaratella SB, Setaro AC, Siegman EC, Subha ZT, Tal R, et al. Pathway of Maternal Serotonin to the Human Embryo and Fetus. Endocrinology. 2018;159(4):1609–29.

67. Thike AA, Chng MJ, Fook-Chong S, and Tan PH. Immunohistochemical expression of hormone receptors in invasive breast carcinoma: correlation of results of H-score with pathological parameters. Pathology. 2001;33(1):21–5.

68. Xu Y, Plazyo O, Romero R, Hassan SS, and Gomez-Lopez N. Isolation of Leukocytes from the Human Maternal-fetal Interface. J Vis Exp. 2015(99):e52863.

69. Bray NL, Pimentel H, Melsted P, and Pachter L. Near-optimal probabilistic RNA-seq quantification. Nat Biotechnol. 2016;34(5):525–7.

70. Durinck S, Spellman PT, Birney E, and Huber W. Mapping identifiers for the integration of genomic datasets with the R/Bioconductor package biomaRt. Nat Protoc. 2009;4(8):1184–91.

71. Soneson C, Love MI, and Robinson MD. Differential analyses for RNA-seq: transcript-level estimates improve gene-level inferences. F1000Res. 2015;4:1521.

72. Love MI, Huber W, and Anders S. Moderated estimation of fold change and dispersion for RNA-seq data with DESeq2. Genome Biol. 2014;15(12):550.

73. Stuart T, Butler A, Hoffman P, Hafemeister C, Papalexi E, Mauck WM, 3rd, et al. Comprehensive Integration of Single-Cell Data. Cell. 2019;177(7):1888–902 e21.

74. Team RC. R: A Language and Environment for Statistical Computing. Vienna, Austria: R Foundation for Statistical Computing; 2020.

75. Hafemeister C, and Satija R. Normalization and variance stabilization of single-cell RNA-seq data using regularized negative binomial regression. Genome Biol. 2019;20(1):296.

76. Zhou Y, Zhou B, Pache L, Chang M, Khodabakhshi AH, Tanaseichuk O, et al. Metascape provides a biologist-oriented resource for the analysis of systems-level datasets. Nat Commun. 2019;10(1):1523.

77. Tang Z, Tadesse S, Norwitz E, Mor G, Abrahams VM, and Guller S. Isolation of hofbauer cells from human term placentas with high yield and purity. Am J Reprod Immunol. 2011;66(4):336–48.

